# One-Seq: A Highly Scalable Sequencing-Based Diagnostic for SARS-CoV-2 and Other Single-Stranded Viruses

**DOI:** 10.1101/2021.04.12.21253357

**Authors:** Mingjie Dai, Wenzhe Ma, Hong Kang, Matthew Sonnett, George M. Church, Marc W. Kirschner

## Abstract

The management of pandemics such as COVID-19 requires highly scalable and sensitive viral diagnostics, together with variant identification. Next-generation sequencing (NGS) has many attractive features for highly multiplexed testing, however current sequencing-based methods are limited in throughput by early processing steps on individual samples (e.g. RNA extraction and PCR amplification). Here we report a new method, “One-Seq”, that eliminates the current bottlenecks in scalability by enabling early pooling of samples, before any extraction or amplification steps. To enable early pooling, we developed a one-pot reaction for efficient reverse transcription (RT) and upfront barcoding in extraction-free clinical samples, and a “protector” strategy in which carefully designed competing oligonucleotides prevent barcode crosstalk and preserve detection of the high dynamic range of viral load in clinical samples. This method is highly sensitive, achieving a limit of detection (LoD) down to 2.5 genome copy equivalent (gce) in contrived RT samples, 10 gce in multiplexed sequencing, and 2-5 gce with multi-primer detection, suggesting an LoD of 200-500 gce/ml for clinical testing. In clinical specimens, One-Seq showed quantitative viral detection against clinical Ct values with 6 logs of linear dynamic range and detection of SARS-CoV-2 positive samples down to ∼360 gce/ml. In addition, One-Seq reports a number of hotspot viral mutations at equal scalability at no extra cost. Scaling up One-Seq would allow a throughput of 100,000-1,000,000 tests per day per single clinical lab, at an estimated amortized reagent cost of $1.5 per test and turn-around time of 7.5-15 hr.

## Introduction

Highly scalable and highly sensitive viral diagnostics (e.g. SARS-CoV-2) are critical for both pandemic response and long-term epidemiological surveillance (*1-3*). During a pandemic, population-wide testing would provide effective control and monitoring of the viral spread, and allow safe return to work. In the long term, regular and population-wide monitoring promises a “bio-weather map” to identify and forecast new viral infection hotspots, preventing the next outbreak. Furthermore, the ability to sequence and identify emerging viral variants (e.g. B.1.1.7, B 1.427), also on the population scale, would allow real-time monitoring of the rate of transmission and pathogenicity, as well as inform public health policies and vaccine development (*4, 5*). Current diagnostic methods fall short of these requirements, as they are limited in either sample processing throughput, testing sensitivity and reliability, or the ability to identify different viral variants.

At present, molecular tests using “gold standard” RT-qPCR in central laboratory facilities have demonstrated high detection sensitivity (down to 200-500 gce/ml) (*6-9*), but are limited in throughput by the requirements of RNA extraction and PCR thermocycling on each sample individually, as well as other liquid handling operations (Fig. S1) (*4*). As a result, it is challenging for most current clinical labs to perform more than 10,000 diagnostic tests per day, even with the help of automation (*10*). By re-purposing large-scale liquid handling and sample automation, up to 100,000 tests per day can be achieved (*11*), but this approach requires heavy upfront capital investment and personnel costs.

Next-generation sequencing (NGS) based methods have long been attractive alternatives to RT-qPCR in two ways: (i) the intrinsic high-throughput readout for multiplexed diagnostics, and (ii) the ability to obtain viral genome sequences for variant identification. In principle the very high throughput readout (up to 10^10^ reads per session, on an Illumina NovaSeq machine) would allow a single testing lab to process up to a million patient samples per day with pooled analysis, if they could avoid the handling of so many individual samples. Since the beginning of the COVID-19 pandemic, several methods for NGS-based multiplexed testing have been proposed and developed (*12-18*). As expected, methods that reported detection sensitivity close to RT-qPCR tests (200-1,000 gce/ml) mostly followed traditional barcoding and sequencing workflows, and required individual RNA extraction and PCR thermocycling steps (Fig. S1) (*12-14, 17*) (or used an extraction-free protocol but with ∼10 fold lower sensitivity (*12, 19*)), which in practice hinders the maximum achievable sample throughput (Fig. 1A). Furthermore, current methods either do not report viral variant information, or perform whole genome sequencing (WGS), which further limits the achievable throughput due to the large number of sequencing reads required.

**Figure 1.**
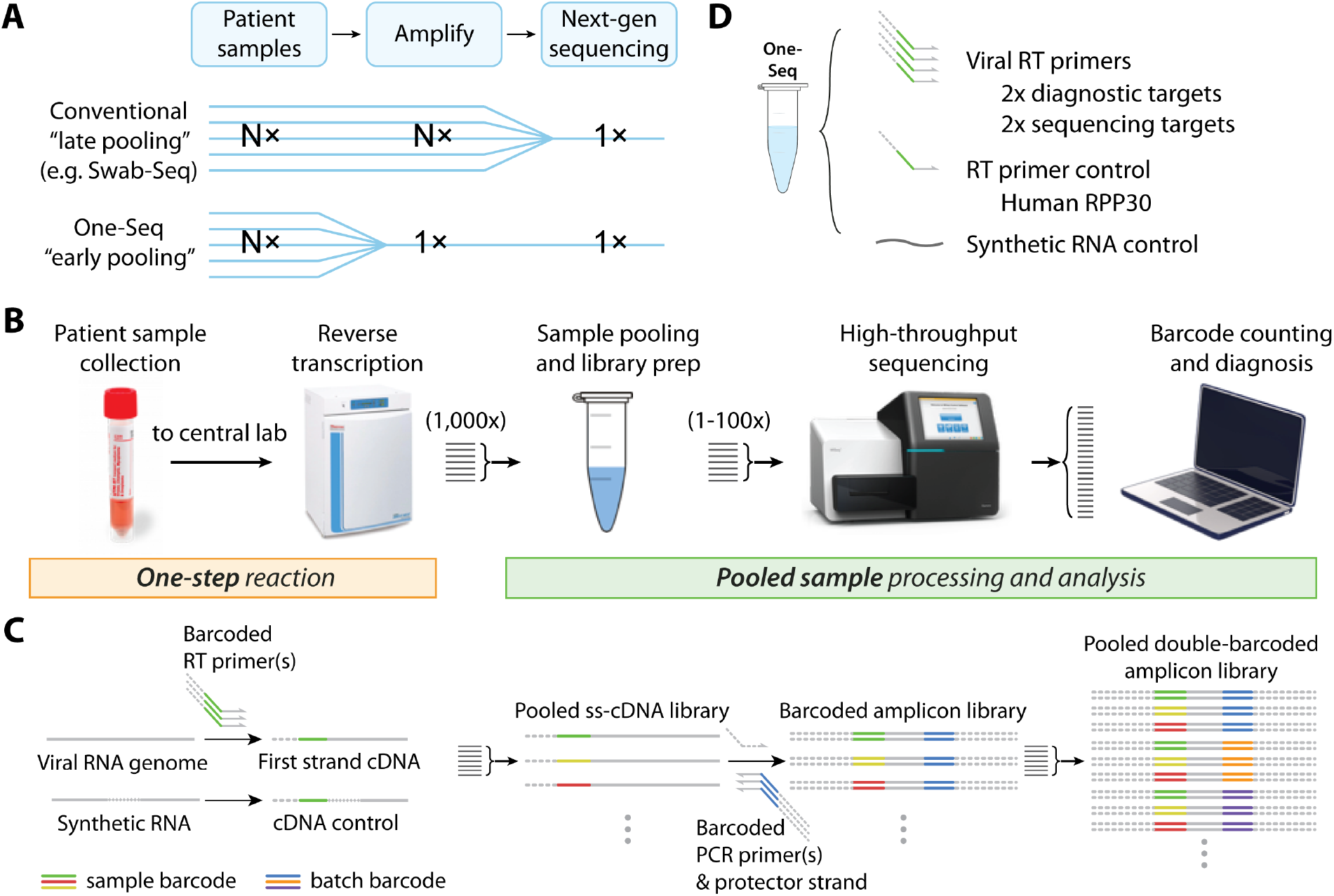
Principle and workflow of One-Seq for highly scalable viral detection and variant identification. (**A**) Illustration of One-Seq “early pooling” strategy in comparison with conventional “late pooling” methods. (**B**) Clinical workflow of One-Seq. Early pooling allows up to 100,000 patient samples to be pooled and analyzed together. (**C**) Molecular workflow of One-Seq. One-Seq uses upfront sample barcoding and a “protector” strategy to enable early sample pooling, and uses a two-stage pooling strategy to support highly scalable testing. (**D**) Illustration of One-Seq reaction components. One-Seq uses multiple RT primers for viral diagnostic and sequencing, one human gene RT primer and one synthetic RNA as positive controls.

To overcome such limitations, we developed a sequencing-based method that achieves high sensitivity, high throughput, and identification of viral variants. To obtain high throughput we implemented a novel “pooling-before-amplification” strategy (Fig. 1A, S1), and developed a new workflow that performs extraction-free, PCR-free, one-step processing from clinical sample to library pooling, thus enabling thousands of patient samples to be processed immediately after arrival at testing centers, all further steps being done in bulk (Fig. 1B). We termed the method “One-step” viral Sequencing, or “One-Seq”.

## Results

The molecular workflow of One-Seq consists of the following steps (Fig. 1C, S2): (1) viral particles from patient samples are lysed and viral RNA is transcribed to a first strand cDNA using a barcoded RT primer, that includes the patient sample barcode and an adaptor for library amplification; (2) barcoded single-stranded cDNAs are pooled (100-1,000 samples) and purified to remove excess RT primers and buffer; (3) second strand cDNA synthesis and PCR library amplification from a common reverse primer and a common forward extension primer are performed together, optionally with a batch barcode on the reverse side; (4) amplicon libraries are cleaned up and normalized, optionally pooled again with different batches, and analyzed by next-generation sequencing. This workflow is further compatible with multiplexed viral detection and sequencing (Fig. 1D), where several strands sharing the same patient barcode but with different RT primer sequences would be mixed together. This multi-primer strategy confers three important benefits: (i) increased detection sensitivity (sensitivity increases linearly with number of primers), (ii) ability to sequence multiple viral loci to enable variant identification, and (iii) simultaneous detection of multiple different viruses (e.g. common cold, flu, SARS-CoV-2 viruses), informing better diagnosis as well as providing a more comprehensive picture for epidemiological surveillance. On top of viral targets, One-Seq further incorporates two positive controls: one against a specially designed synthetic RNA fragment that shares the same RT primer as one of the viral targets but has different sequence, and another against the human RPP30 gene (Fig. 1D).

To implement this workflow, we faced two critical challenges. First, our one-step, extraction-free reaction has to perform three tasks simultaneously: viral lysis and release of viral RNA, efficient reverse transcription that allows high-sensitivity viral detection, and preservation of patient samples at room temperature for up to 24 hr during sample collection and transport to the central lab. Second, by performing pooling before amplification, the library amplification reaction must faithfully preserve the high dynamic range of viral load known to exist in clinical samples (up to a 10^6-7^-fold range) (*20*), and at the same time achieve high detection sensitivity. In particular, the method needs to stringently avoid any barcode crosstalk that may arise from amplification and sequencing steps, as this would result in false positive diagnoses.

### Development of a one-pot reaction for efficient viral reverse transcription and sample preservation

#### An optimized RT reaction system allows for sensitive RNA detection from extraction-free samples

Our first goal was to develop an extraction-free and high-sensitivity method for viral lysis and reverse transcription (RT), in the presence of potential inhibitors in patient samples (e.g. NP swab or saliva). We reasoned that, since reverse transcriptases are in general more resistant to inhibitors than thermostable polymerases, there might be an unappreciated advantage in separating the RT and PCR steps in the traditional RT-PCR workflow, since this would allow more flexibility in formulating the RT reaction mix. To test this hypothesis and assay RT efficiency in the presence of inhibitors, we prepared contrived standard samples with human saliva collected from COVID-19 negative donors and viral RNA spike-in (synthetic RNA fragment generated by in vitro transcription (IVT), or full-length RNA genome from Twist BioSciences). We first compared RNA protection effects of different RNAse inhibitors and found Murine (New England Biolabs) and RNAsin (Promega) provided the best and similar protection at 25°C to 50°C. We then compared RT efficiency of various reverse transcriptases in saliva-containing samples (Fig. S3), using qPCR readout with CDC’s RT-PCR primer and probe set (*21*). Our results showed that SuperScript IV reverse transcriptase can detect 3 molecules of synthetic RNA in the presence of human saliva, making it a promising candidate for the development of an efficient, extraction-free reaction.

We next prepared contrived clinical samples using pooled COVID-19 negative remnant clinical specimens (nasopharyngeal (NP) swab in viral transport medium (VTM), N=15), with spiked-in inactivated virus standard (heat-inactivated SARS-CoV-2 from ATCC, or AccuPlex SARS-CoV-2 verification panel from SeraCare) (Fig. 2A). In contrast to a “naked” RNA spike-in, these inactivated virus samples allowed us to assay the efficiency of viral lysis in patient samples.

**Figure 2.**
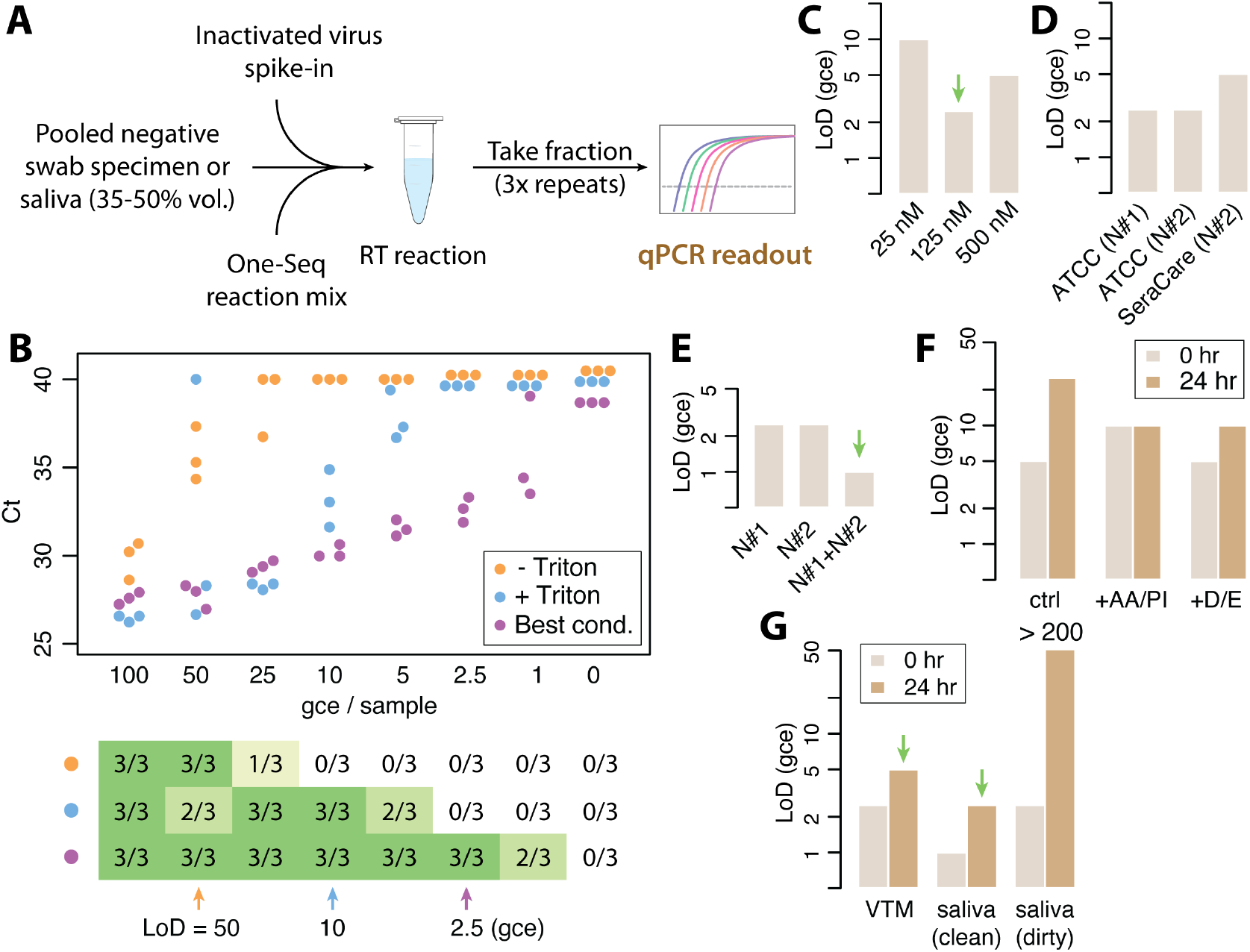
Development of an extraction-free, one-pot reaction for efficient viral reverse transcription and sample preservation. (**A**) Schematic of RT efficiency test in contrived clinical samples, using pooled negative specimen and inactivated virus spike-in. (**B**) Example of the RT sensitivity test, top, Ct values (3x repeats) plotted against different viral loads, in genome copy equivalent (gce), bottom, detection rate and limit of detection (LoD) determination. (**C**-**E**) RT sensitivity test under different conditions. (**C**) Comparison of different RT primer concentrations. (**D**) Comparison of different RT primers and validation with different virus reference standards. (**E**) comparison of single-vs dual-primer detection. (**F**-**G**) Effect of sample preservation buffer after incubation for 0 or 24 hr at room temperature in clean reaction buffer (**F**) or contrived patient samples (**G**). AA: antibiotic and antimycotic, PI: protease inhibitor, D: DTT, E: EDTA, VTM: viral transport medium.

To assay the analytical sensitivity of the RT reaction, we prepared a roughly 2x dilution series of inactivated virus standard (ATCC) in contrived clinical samples, ranging from 100 genome copy equivalent (gce) to less than 1 gce per reaction. The RT product was assayed by qPCR in triplicate (Fig. 2B). We found that the RT samples indeed showed a significant inhibitory effect on PCR amplification, consistent with our earlier hypothesis; PCR efficiency is restored only after a 40-80x dilution.

To optimize viral lysis, we tested the effect of detergents, which have been previously mentioned to help with viral lysis and RNA release (*22, 23*). We found that, the addition of mild detergent (Triton X-100) improved the detection sensitivity by ∼5x from extraction-free viral samples, from a limit of detection (LoD) = 50 gce to 10 gce (3/3 detection) (Fig. 2B). We designed two RT primers against the viral N gene, optimizing thermodynamic parameters and avoiding regions with significant sequence variance or homology to other related viruses (Table S1). After optimizing for primer concentration (Fig. 2C, S4), we showed that with detergent both primers could achieve an LoD = 2.5 gce, close to theoretical maximum sensitivity (Fig. 2B,D, S4). We further verified our detection limit with a different source of viral reference standard (SeraCare), and obtained similar results (Fig. 2D, S4).

Multiplexed RT with multiple primers offers the potential of increased detection sensitivity. We tested this effect with our two N-gene-targeting primers in contrived clinical samples. Indeed, we observed a roughly 2-fold higher detection sensitivity (LoD = 1 gce) when signals from both primers were considered (Fig. 2E, S4). We note that, since both primers target different genomic loci (separated by ∼800 nt), the detection of these loci can be considered as independent events and thus it is possible to obtain LoD values of less than 2 molecular copies.

#### One-Seq sample stabilization buffer preserves clinical samples and allows sensitive detection after 24 hr incubation at room temperature

We further developed a one-pot reaction system that can also stabilize patient samples for up to 24 hr at room temperature, which allows for a delay between sample collection and transport to central testing lab. To work out the parameters we started with contrived saliva samples with synthetic RNA spike-in (IVT), and screened a list of potential candidates for their sample preserving effect, including antibiotics and antimycotics, protease inhibitors, reducing agents and metal chelating agents. We further grouped the promising candidates into RNA-preserving (EDTA and DTT) and RT enzyme-preserving (antibiotic and antimycotic, protease inhibitor) factors, then tested their effects in contrived clinical VTM samples prepared as above, with inactivated virus spike-in. After 24 hr incubation at room temperature, both groups individually improved RT efficiency by roughly 2-fold (Fig. 2F, S5); together they improved detection sensitivity significantly (from LoD = 25 gce to 5 gce), only a 2-fold reduction compared with unincubated (0 hr) control (Fig. 2G, S6).

We also tested our sample stabilization buffer in contrived saliva samples (Fig. 2G, S6). For this test, we compared saliva specimens from COVID-19 negative donors collected with or without careful mouth rinsing beforehand (denoted as “clean” and “dirty” saliva samples). We pooled saliva specimens for both cases (N=4 and N=9, respectively), and spiked in inactivated viral standard (Fig. 2A). Without room temperature incubation, both contrived saliva samples allowed highly sensitive detection (LoD <= 2.5 gce). After 24 hr incubation, we could still detect viral RNA with high sensitivity (LoD = 2.5 gce) in the “clean” saliva sample, suggesting our sample stabilization buffer successfully preserved the viral genetic material without significant degradation (Fig. 2G, S6). On the other hand, all signal was lost in samples containing “dirty” saliva (with visible food particles and other suspended debris), likely due to the degrading effect of food residues and microbes present in these samples.

### Development of a “pooling-before-amplification” workflow for high sensitivity and high dynamic range multiplexed sequencing

#### Barcode selection and cDNA purification allows efficient amplification after sample pooling

Our second goal was to develop a “pooling-before-amplification” workflow for sample pooling and PCR library amplification that not only maintains the high detection sensitivity, but also preserves high sample dynamic range and allows quantitative report of viral load in patient samples.

We first designed a set of PCR primers for efficient library amplification (Table S1). For each RT target, we designed several different reverse primers and selected the best for library amplification efficiency by qPCR, and band purity by gel electrophoresis. We wished to design a large set of distinct sample barcodes that are error-tolerant and color-balanced for Illumina’s sequencing machines, and started with the IDT for Illumina unique dual (UD) index set (384 dual index pairs) that had been pre-optimized. We concatenated both indices, and expanded it to 960 unique barcodes by inserting three blocks of sequence tags (Fig. 3A). This method ensures a minimum Hamming distance of 12 between any two barcodes, and thus is tolerant to up to 6 nucleotide substitutions and resistant to even a high level of polymerase and sequencing errors. To select for barcodes that have low secondary structure and are compatible with our workflow, we synthesized barcoded RT primers with all 960 barcodes (Table S2) and pooled 10x 96-well plates of contrived samples using synthetic viral RNA spike-in. After pooled amplification and sequencing, those barcodes that produced read counts higher than a set threshold were selected and used for subsequent tests (Fig. 3B, S7A).

**Figure 3.**
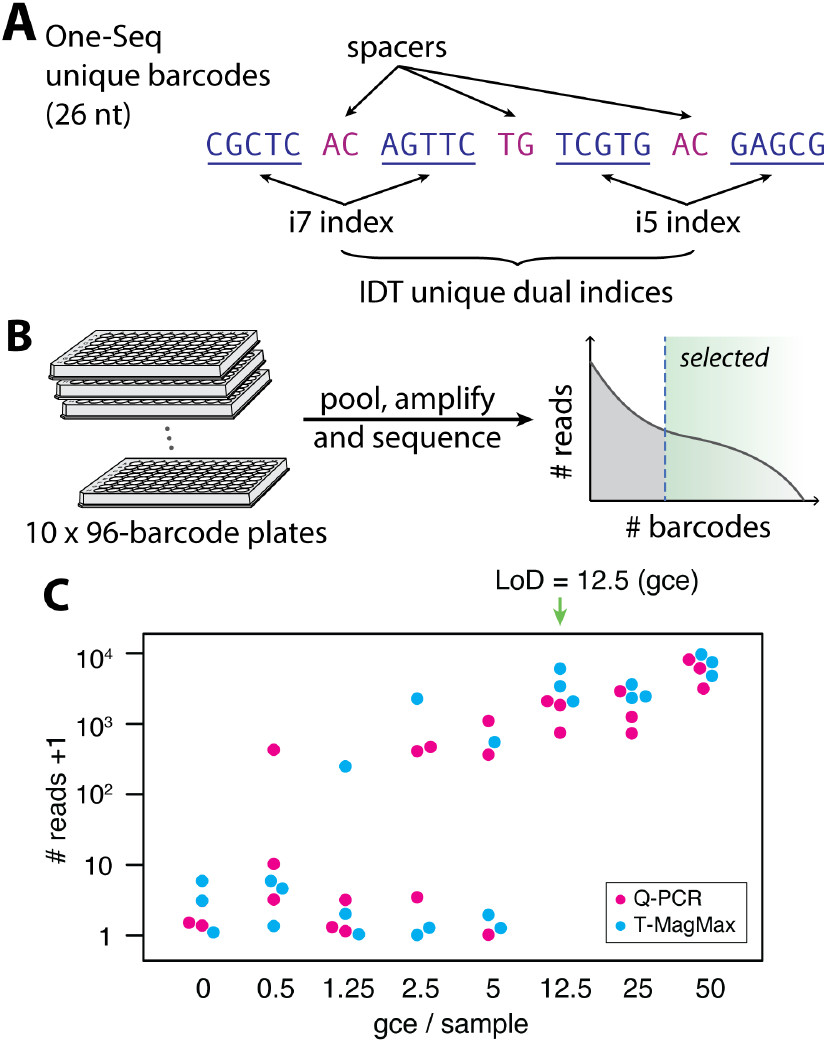
Barcode design and multiplexed sequencing sensitivity test. (**A**) Schematic of unique sample barcode construction. (**B**) Schematic for 960 sample pooling and barcode selection. (**C**) Example multiplexed sequencing sensitivity test and LoD determination, plotted as sequencing read count +1 against expected viral load. cDNA purification allows efficient library amplification after pooling.

We performed a preliminary test on amplification efficiency and dynamic range on these selected barcodes, with a 10x dilution series (Fig. S7B). For high-load samples, we observed a linear response with a mediocre dynamic range of ∼10^4^; the detection sensitivity was also quite low, likely due to PCR inhibitors present in pooled RT samples. To improve PCR amplification efficiency, we performed spinning column cDNA purification after sample pooling. This step also had the added benefit of reducing sample volume to a manageable level, after pooling a large number of patient samples. After cDNA purification and using 96 selected high-quality barcodes (Table S3), our results showed an LoD = 12 gce (Fig. 3C), which is about 5-fold lower than the qPCR readout, suggesting some degree of sample loss and degradation during the cDNA purification, library amplification and sequencing steps.

#### Dynamic strand displacement using a “protector” oligonucleotide effectively suppresses barcode crosstalk and preserves sample dynamic range

Suppressing off-target barcode crosstalk and preserving high sample dynamic range are critical for faithful COVID-19 diagnostics, as clinical samples have been shown to exhibit a large dynamic range (up to 10^6-7^) of detectable viral load (*8, 20*), and any barcode mis-assignment could result in false positive diagnoses. We first assayed the degree of barcode crosstalk in our workflow by pooling 1 or 10 barcoded RT samples prepared with high spiked-in viral load together with 95 or 86 negative samples with other barcodes, and tallied sequencing reads carrying any of the off-target barcodes (Fig. 4A). Without any special treatment, our results showed a 0.1% barcode crosstalk on average, resulting in an upper limit of 3 logs of detectable sample dynamic range (Fig. 4B), much lower than what is required for faithful COVID-19 diagnostics when a high-load sample is present.

**Figure 4.**
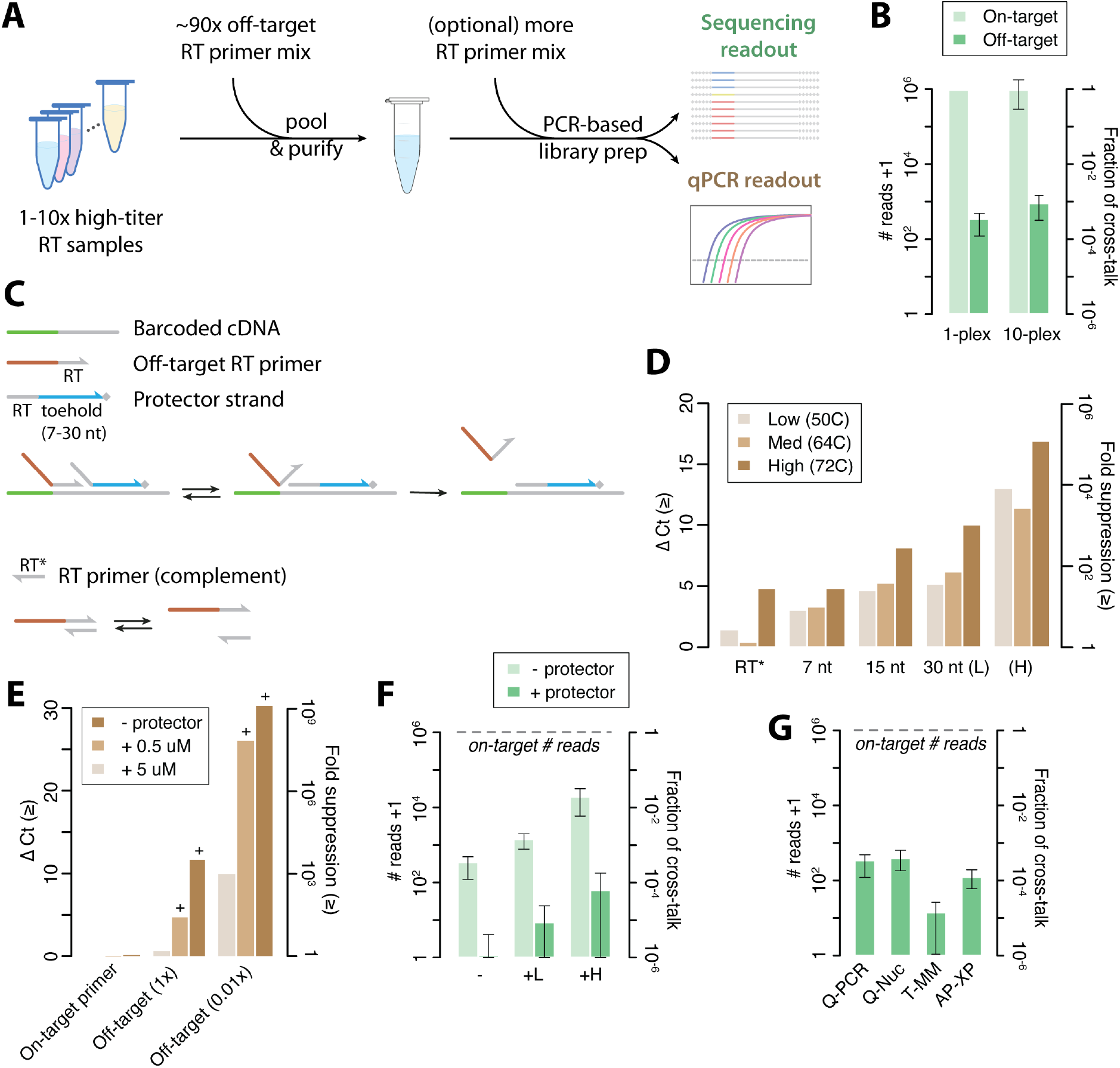
Development of a “protector” strategy that suppresses barcode crosstalk and preserves large sample dynamic range. (**A**) Schematic for barcode crosstalk and dynamic range test by qPCR and multiplexed sequencing readout. (**B**) On-target and off-target sequencing read counts and fraction of crosstalk without using the protector strategy. (**C**) Schematic for two approaches to suppress barcode crosstalk: top, dynamic strand displacement with a protector strand, bottom, a naïve approach with complementary strand hybridization. (**D**-**E**) Crosstalk and dynamic range test with 1 on-target amplicon and 1 off-target primer, assayed by qPCR under different conditions. (≥) indicates lower bounds. (**D**) Effect of protector strand design and annealing temperature. (**E**) Effect of off-target primer and protector strand concentration. Bars with a plus sign on top are samples that showed abnormal melt curve, suggesting a higher degree of suppression than measured. (**F**-**G**) Crosstalk and dynamic range test with 1 high-load sample and 95 off-target RT primers, assayed by multiplexed sequencing under different conditions. (**F**) Effect of supplementing extra off-target primers (+L, low amount, +H, high amount), with and without using the protector strategy. (**G**) Comparison of different cDNA purification methods. Q-PCR, QIAquick PCR purification kit (QIAGEN), Q-Nuc, QIAquick nucleotide removal kit (QIAGEN), T-MM, MagMax viral/pathogen nucleic acid isolation kit (ThermoFisher), AP-XP, AmPure XP PCR purification beads (Beckman Coulter).

A major source of barcode crosstalk in the traditional, “pooling-after-amplification” workflow is from cross-hybridization of excess library adapters during the cluster amplification process, which then produces mis-barcoded transcripts (*24*). We hypothesized that, a similar mechanism with cross-hybridized excess RT primers during the library amplification step could account for the main source of the 0.1% barcode crosstalk we observed. Unfortunately, the traditional method for minimizing crosstalk using unique dual indices is not compatible with a “pooling-before-amplification” strategy. However, we thought it should be possible to devise a strategy that reduces such crosstalk by suppressing cross-hybridization of excess RT primers (Fig. 4C, top). To do this we designed a single-stranded “protector” oligonucleotide that comprises the RT primer (without barcode), an extended sequence complementary to the viral genome downstream, and a polymerase blocker to prevent off-target amplification. By the principle of dynamic strand displacement (*25*), the extended sequence functions as a toehold and provides stable binding of the protector strand to the cDNA, which would then compete off any off-target RT primer from cross-hybridization (Fig. 4C, top).

We first performed a simple test of this protector strategy, using a short DNA amplicon together with an off-target barcoded RT primer, using qPCR as the readout. We compared a few different protector strand designs, including a naïve approach using simply the complement of the RT primer sequence (Fig. 4C, bottom). We observed that the protector strand significantly reduced off-target PCR amplification; longer toehold lengths (up to 30 nt) provided more stable binding, leading to more effective suppression (Fig. 4D). Increasing protector strand concentration and raising annealing temperature also each improved the suppression effect, as both favor the binding of the protector strand compared to that of the off-target primer. Under optimized conditions, our results showed up to 10^5^-fold suppression of off-target amplification. We further tested the effect of RT primer concentration (Fig. 4E). Our results showed that, lowering RT primer concentration by 100x alone can reduce barcode crosstalk by 1,000-fold; and an overall 10^9^-fold suppression can be achieved when used in combination with the protector strand.

We next tested the protector strategy in multiplexed sequencing settings and in contrived clinical samples, following similar test design as used previously (1-10 high-load sample along with ∼90 off-target barcodes) (Fig.4F, S8). Using the protector strategy significantly reduced the level of barcode crosstalk from 0.03% to 0.0001% (i.e. 300-fold reduction). We then stress tested the system by supplementing extra off-target RT primer mix into the PCR reaction (Fig.4F). Without adding the protector strand, we observed a significantly higher barcode crosstalk (0.1%-6%), confirming our earlier hypothesis; with the protector strand, the crosstalk level is again significantly suppressed (0.001-0.01%). To further reduce barcode crosstalk, we compared the effect of RT primer removal by several cDNA purification methods (Fig. 4G, S8). Our results showed that bead-based purification methods (e.g. Thermo MagMax kit) produced a lower level of barcode crosstalk (0.001%) compared to spin column based ones (e.g. QIAquick PCR purification kit), likely due to a sharper size selection cut-off. Since our spiked-in samples has a very high viral load (equivalent to 2×10^9^ gce/ul in patient sample, or Ct=12), we expect a much lower level of barcode crosstalk in practical scenarios, allowing for a dynamic range of 10^6-7^, fulfilling the requirement for faithful SARS-CoV-2 detection in patient samples with our highly multiplexed method.

### Validation of One-Seq in clinical samples

We validated the performance of our method in SARS-CoV-2 positive clinical samples (Fig. 5A). To mimic conditions where samples are directly collected into One-Seq reaction, we purchased remnant clinical NP swab samples that had not been heat-inactivated, and compared samples collected in several different viral transport media. We found that samples collected in most commonly used viral transport media were compatible with One-Seq reaction buffer. Only Hologic Aptima swab samples were incompatible, generating snow-like aggregates, most likely due to precipitation of lauryl sulfate in the Aptima buffer with potassium ions in our buffer.

**Figure 5.**
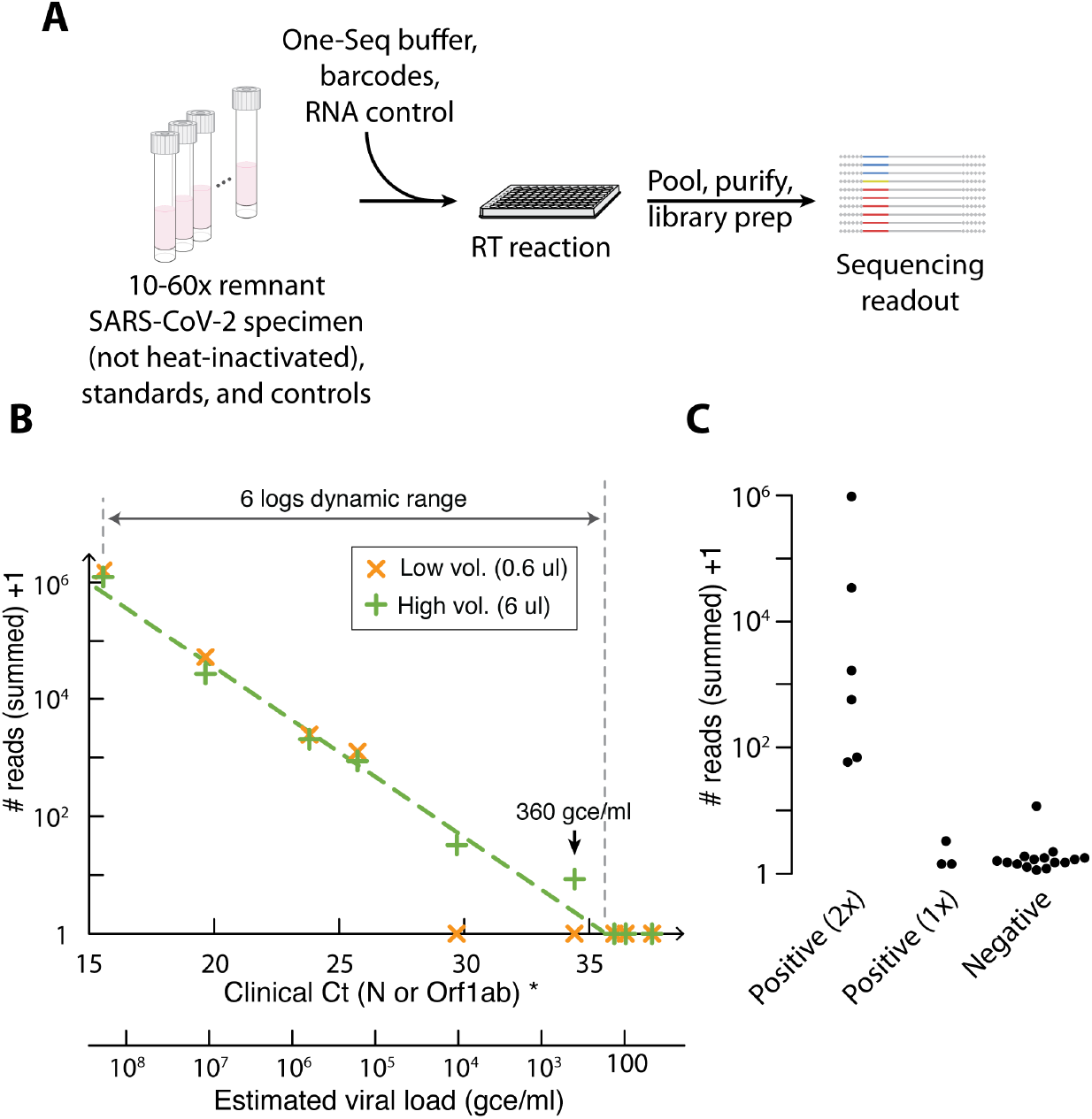
Validation of One-Seq on clinical SARS-CoV-2 specimens. (**A**) Schematic of One-Seq test with remnant clinical specimens. (**B**) Example of One-Seq testing results, plotted as One-Seq sequencing read counts (summed) +1 vs clinical Ct values by RT-qPCR and estimated viral load (calculated according to manufacturer’s specification). One-Seq results showed 6 logs of linear dynamic range with respect to patient viral load, and correctly detected samples down to 360 gce/ml. (^*^) For samples without a valid Ct(N) value, Ct(orf1ab) is used for plotting. (**C**) Beeswarm plot of One-Seq results for positive (2x), (1x) and negative clinical samples, where positive (2x) refers to samples for which clinical RT-qPCR test returned positive results for both N and orf1ab amplicons, and positive (1x) refers to samples for which only one of the two amplicons were clinically detected (and Ct>36).

To test the detection sensitivity as well as dynamic range of our method, we chose a set of representative COVID-19 positive samples (NP swab in VTM) that spanned a wide range of clinical Ct values (from 15 to 38), and subjected them to the One-Seq workflow. For this test, we mixed three distinct barcodes together for each sample and summed their sequencing reads, to maximize the sensitivity and robustness of detection. We first investigated the detection sensitivity of One-Seq and its dependence on input sample volume (Fig. 5B). As expected, higher sample volume allowed higher detection sensitivity. With only 6 ul per sample input, our method correctly reported the presence of SARS-CoV-2 RNA in all samples with a clinically determined Ct value <35, and no false positives.

According to the manufacturer’s specification (*26*), the lowest sample concentration we detected was at 360 gce/ul (Ct = 34.39), suggesting that One-Seq can detect clinical samples with viral load in the 200-500 gce/ul range, using a single amplicon. We also observed a linear correspondence between our reported sequencing reads and estimated viral load (calculated from clinical Ct values), over the entire range of Ct values (from 15 to 35), demonstrating that One-Seq faithfully reports viral load in a quantitative manner over 6 logs of dynamic range (Fig. 5B). We observed a slight ratio compression in our sequencing reads, possibly resulting from a decreased RT reaction efficiency in high-load samples, due to the constraints in RT primers and enzymes available. We then performed a second test with both COVID-19 positive and negative samples (NP swab in VTM, total N=28), and observed a clear separation between these samples (Fig. 5C).

In this test, there were three clinically determined positive samples that were not detected. Notably all three had only one of the two targets detected by RT-qPCR (i.e. either the N gene or orf1ab gene target was not detected), and they all had Ct values >36 for the detected target. We believe that if these samples were indeed positive, they were likely missed by our test due to the small sample volume (6 ul) used in this test as compared to a typical RT-qPCR test (300 ul or more) (*10, 27, 28*); further increasing sample volume is expected to improve the detection sensitivity.

### Multi-primer detection and variant sequencing

Simultaneous detection using multiple RT primers potentially allows multi-locus, multi-virus diagnostics, and with increased viral detection sensitivity. Furthermore, if the RT primers are designed to be in close proximity to mutation hotspots (Fig. 6A), it is possible to obtain extra viral sequence information and allow variant identification, without significantly increasing the test turn-around time. The developments in the current pandemic suggested that a very useful application of One-Seq would be for surveillance of viral variants or simultaneous detection of multiple viruses.

**Figure 6.**
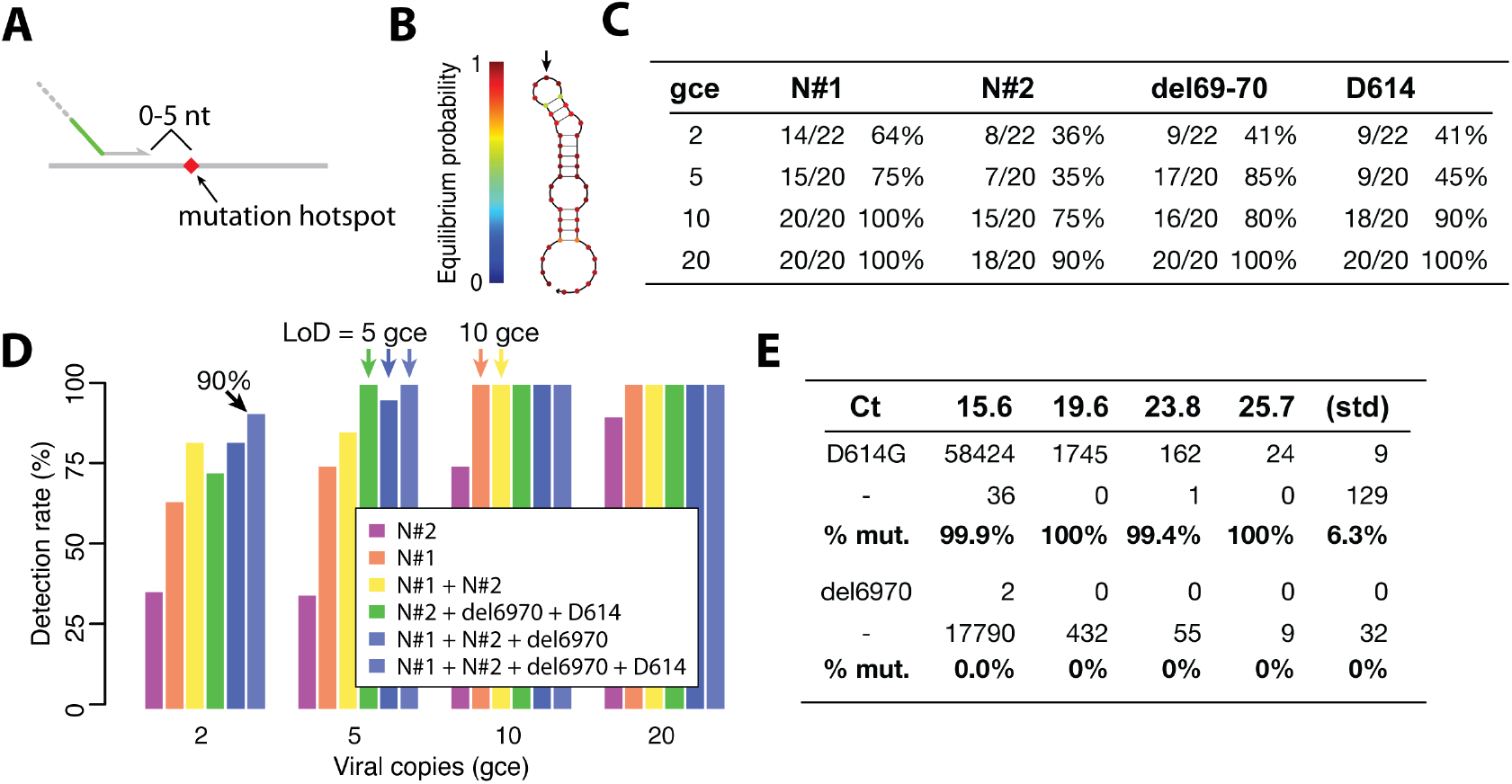
Multi-primer testing and variant sequencing. (**A**) Schematic for RT primer design targeting a viral mutation hotspot. (**B**) Example of strong local secondary structure in the viral genome that prevents efficient RT. Arrow indicates the mutated nucleotide. (**C**) Confirmatory sensitivity test results in contrived clinical samples for all four primer pairs (two in N gene and two in S gene for mutation sequencing) designed for One-Seq. (**D**) Comparison of detection sensitivity with different number and combination of primers. Combining more primers allows higher detection sensitivity, down to LoD = 2-5 gce with all four primers. (**E**) Viral sequencing showed all positive clinical samples we tested had D614G mutation, but none had the del6970 mutation, suggesting they were not related to the B.1.1.7 variant. Raw sequencing reads from four exemplary specimens as well as the virus standard sample (ATCC) were listed.

We designed RT primers targeting several characteristic mutations of the recently reported variant B.1.1.7, in the viral S gene, including del69-70, del144, N501Y, D614G and A701V, and used dye-based qPCR to assay their RT efficiency. We found that it was not always easy to design good RT primers in close proximity to the target mutations, likely due to the presence of strong local secondary structure (Fig. 6B). As a result, our first batch of primer designs only yielded two good candidates with high RT sensitivity (LoD ≤5 gce) (Fig. S9). We performed preliminary sensitivity test for these two primers in contrived clinical samples and in 96x multiplexed format, and our results suggested limits of detection of 10-30 gce for both primers.

We performed in silico analysis for primer inclusivity and specificity for all primer pairs we designed, following FDA guidelines. We found that all primers aligned to all available SARS-CoV-2 genome sequences in NCBI database (98,765 sequences, retrieved on Mar 9^th^ 2021) with at most 1 base mismatch, and 7 out of the 8 primers showed exact match to >99.4% of all sequences (Table S4). Since One-Seq performs RT and PCR in separate steps, we only performed cross-reactivity analysis on RT primers. All four RT primers showed no significant (>80%) homology to genome sequences of common respiratory flora and other related viruses (Table S5). In addition, One-Seq reads a short sequence into the viral genome, providing highly specific viral detection.

We next performed confirmatory clinical sensitivity test for all primer pairs) we designed (4 in total), in a similar 96x multiplexed format, in both single- and multi-primer settings (Fig. S10). For this test, we used only one unique barcode per sample. Our results showed that, in single-primer tests, all four primer pairs had an LoD = 20 gce by the 95% detection rate cut-off (Fig. S10A-C), confirming the results from preliminary studies. In multi-primer tests, three of the four primer pairs still performed well and showed an LoD of 10-20 gce (95% cut-off; all four LoD ≤20 when using a 90% cut-off), and primer N#1 showed an even high sensitivity at LoD = 10 gce (95% cut-off) (Fig. 6C, S10D,E). These results suggest that multiplexed RT and library amplification can work well and there is no significant interference between the designed primers. We also tested if the use of multiple primers would further improve detection sensitivity. Indeed, we observed a higher detection rate as more primers are used (Fig. 6D). If all four primers were used, we obtained an LoD of 5 gce (95% cut-off; 2 gce at 90% cut-off).

For a 4-primer multiplexed test with a 10 ul patient sample intake, this result translates to an LoD of 200-500 gce/ml in clinical samples, approaching the detection limit of RT-qPCR tests. Further increasing sample input volume, or using more primers in parallel would both further increase the detection sensitivity in a linear fashion, e.g. taking 300 ul specimen (typical for RT-qPCR tests (*10, 27, 28*)) would in principle allow an LoD down to 5-10 gce/ml.

Finally, we tested One-Seq with multi-primer detection in clinical samples in a 96x multiplexed format, consisting of 56 clinical samples (two repeats of 28 specimens), 24 contrived standards, and 16 no target controls (Fig. S11). We used all four RT primers designed above, two for diagnostics and target the N gene, two for mutation sequencing and target the S gene. Using only 5 ul sample volume, One-Seq correctly reported the low viral load sample (360 gce/ul) in both repeats, and again exhibited a linear dynamic range of ∼10^6^, allowing quantitative reporting of viral load. Due to an unexpected manufacturer delay, we have not been able to use all primers together to test further improvement of detection sensitivity. As we looked into viral sequences from the two mutation-targeting primers in the S gene (Fig. 6E), we found that the D614G mutation was present in all positive clinical samples we tested, but not the inactivated virus standard (isolate USA-WA1/2020, Jan 2020), suggesting that the D614G mutation was already prevalent in July 2020, when this batch of samples was originally collected. Consistent with our expectation, we found no evidence of the del6970 mutation, suggesting that none of these samples were related to the later discovered B.1.1.7 variant.

## Discussion

We report here a new method for viral RNA molecular diagnostics (e.g. SARS-CoV-2) that allows highly scalable central lab testing, achieves high detection sensitivity, and provides sequence information at targeted mutation hotspots, allowing for viral variant identification. One-Seq can take unextracted samples, either inactivated or not, and reach a high detection sensitivity down to 10 gce by multiplexed sequencing using a single primer, and down to 2-5 gce for multi-primer detection with four primers. Assuming 10 ul sample intake, this is equivalent to a viral load of 200-500 gce/ml in unextracted patient sample, approaching the maximum sensitivity of extraction-based RT-qPCR assays. Scaling up sample volume should further improve the detection sensitivity linearly. In clinical samples, One-Seq quantitatively reports patient viral load, preserves 6 logs of linear dynamic range of viral load (estimated from clinical Ct values), and detected SARS-CoV-2 positive samples down to 360 gce/ml in viral load. One-Seq further reports sequences at a number of viral mutation hotspots, allowing for viral diagnostics and variant identification in a single test, at equal scalability and no extra cost.

One-Seq can be used with a two-stage barcoding and pooling strategy to test a large number (e.g. 100,000) of patient specimens, without the need to design and manufacture an equally large number of distinct barcodes (Fig. 1B,C). To implement this strategy, patient specimens can be collected in different batches (e.g. by local community, organization, or department) up to a certain size (e.g. 1,000 samples per batch). Samples in each batch will be pooled, purified and processed together. Each batch will then be barcoded on the reverse side during the library amplification step, after which a number of batches will be pooled together for multiplexed sequencing. This two-stage barcoding strategy provides two benefits. First, it significantly reduces the overhead in barcode design, manufacturing and regulatory approval. Second, it allows the method to be adapted and applied to different application scenarios, for example in an isolated environment (e.g. a cruise ship) where only a limited number of individuals needs to be tested regularly. In such a scenario, One-Seq can be adapted to use the same barcode set but with less second-stage pooling, and sequenced on a lower-throughput machine (e.g. Illumina NextSeq 550).

One-Seq is highly scalable, cost-effective, with a fast turn-around (Table 1). Using a high output Illumina sequencer such as the NovaSeq 6000, we estimated a maximum sample throughput of 250,000-400,000 per day per machine, making possible an overall throughput of up to 1,000,000 tests per day in a single clinical lab, with the help of three sequencers. Further increase in sample throughput as well as cost reduction are possible with other sequencing modalities, e.g. Oxford Nanopore PromethION 48 allows up to 5x lower sequencing reagent cost, and up to 450,000 tests per day at comparable capital cost, although we have not validated this approach in the current study (Table 1). Depending on the sequencer model used and whether batch pooling and viral sequencing are desired, One-Seq sample turn-around time (TAT) ranges from a minimum of 7.5 hr (for a single batch on a MiSeq, without viral sequencing) to a maximum of 14.5 hr (for batch pooling on a NovaSeq 6000, with viral sequencing), allowing for diagnostic results to be available within 24 hr of sample collection or drop-off (Table S6). The cost per sample for One-Seq also scales favorably for highly multiplexed settings. We estimated that, while at relatively small scale (e.g. 200 samples per run on a MiSeq) and using off-the-shelf reagents, the cost is at $9.5 per test; at large scale (e.g. 100,000 samples per run on a NovaSeq 6000) sequencing reagent cost is reduced to <$0.2 per sample, and mass production is expected to lower enzyme and reagent cost by 70% or more, bringing the total cost down to $1.5 (Table S7). Due to the minimum sample processing needed for the One-Seq workflow, we expect the consumable cost (e.g. tips, tubes) will also be considerably lower, making the total cost per test lower than that for current RT-qPCR or sequencing-based methods. In addition to scalability, One-Seq also shows superior performance in comparison with other methods, and offers high detection sensitivity (down to LoD = 200-500 gce/ml), and ability to test unextracted clinical samples (Table 2). We believe that One-Seq offers a technically and economically viable solution for highly scalable testing on a population scale.

**Table 1.** Key performance characteristics for scalable diagnostics with One-Seq.

| One-Seq specification | MiSeq | NextSeq 550 | NovaSeq 6000 | PromethION 48* |
| --- | --- | --- | --- | --- |
| Max. samples per run ** | 200 | 4,000 | 100,000 | 150,000 (8 hr) |
| Max. samples per day<br>(diagnostics only) | 1,200 | 16,000 | 400,000 | 450,000 |
| Max. samples per day<br>(diagnostics & sequencing) | 800 | 12,000 | 250,000 |  |
| Sequencing cost | \$ 4.2 | \$ 0.4 | \$ 0.12 | <\$ 0.1 |
| Cost per sample (off-the-shelf ***) | \$ 9.5 | \$ 5.7 | \$ 5.4 | \$ 5.4 |
| Cost per sample (mass produced ***) | \$ 5.6 | \$ 1.8 | \$ 1.5 | \$ 1.5 |
| Turn-around time | 7.5-10.5 hr | 9-12.5 hr | 10-14.5 hr | 12 hr |

\* Scaled (2x) to match capital cost as one NovaSeq 6000 sequencer
\*\* Assuming an amortized $1 \times 10^5$ sequencing reads per sample, this allows for up to 3-5% of high viral load samples.
\*\*\* All cost estimated assuming 10 ul patient sample volume. For cost with mass production, see Table S7 for details.

**Table 2.** Performance comparison between One-Seq and other methods (*6, 7, 9, 12, 14*)

| METHOD | RT-qPCR |  | COVIDSeq | Swab-Seq |  | LamPORE | One-Seq |
| --- | --- | --- | --- | --- | --- | --- | --- |
|  | With RNA extraction | Without extraction |  | With RNA extraction | Without extraction |  |  |
| Throughput-limiting Step | RNA extraction / RT-PCR | RT-PCR | Sequencing | RNA extraction | PCR ** | RNA extraction | Decapping / Sequencing |
| Max. samples per day per limiting instrument * | 1,600 | 1,600 | 1,000 | 4,800 | 6,400 ** | 4,800 | 250,000-450,000 |
| Sensitivity (gce / ml) (self-reported) | 20-500 | 2,000+ | 1,000 | 250 | 1,000-3,000 | 20-200 | 1,000-2,000 <sup>†</sup><br>200-500 <sup>††</sup> |
| Sensitivity (gce / ml) (FDA reference panel test) | 180-18,000 | - | 5,400 | - | - | - | - |
| Viral sequencing capability | - |  | Whole genome | Targeted (2x) |  | Targeted (multiple) | Targeted (multiple) |
| Reagent cost (amortized) | \$ 3-6 | | \$20 | \$ 2-4 *** | | \$ 3-6 *** | \$ 6.8 <sup>†</sup><br>\$ 1.5 <sup>††</sup> |
| Turn-around time | 2-4 hr |  | 12-24 hr | 8 hr |  | 6 hr *** | 7.5-15 hr |

\* For RNA extraction or RT-PCR limited tests, throughput is estimated assuming sample processing in 96-well formats, and under the assumption that RNA extraction takes 0.5 hr, and PCR thermocycling takes 1.5 hr.
\*\* PCR throughput is estimated using 384-well plates. Further scaling up is possible with high-throughput thermocycler, e.g. Hydrocycler2 to 16x, if assuming no significant delay in sample processing and plate aliquoting.
\*\*\* Estimated values.
<sup>†</sup> Demonstrated performance in the current work, using 5 ul sample, off-the-shelf reagents, and run on a MiSeq.
<sup>††</sup> Projected performance using 10 ul sample, four primers and run on a NovaSeq 6000.

One-Seq also allows detection of viral hotspot mutations and monitoring of their transmission dynamics (Table 2). This is especially important as certain mutations could convey higher transmission rate or pathogenicity (e.g. B.1.1.7 (*29*)), or evasion from immunity induced by vaccination or prior infection (e.g. E484K) (*30, 31*). It has been increasingly appreciated that identifying and tracking viral variants is as critical as diagnostic screening, and sequencing remains the only method available for effective variant identification (*5*). Current whole-genome sequencing (WGS) methods (e.g. Illumina COVIDSeq (*13*)) typically require 50-100x sequencing reads for the same sample, and are further bottlenecked in throughput by the PCR-limited sample preparation steps. In contrast, One-Seq uses targeted sequencing that requires much fewer reads per sample and allows much higher scalability and lower amortized cost. Therefore, we believe One-Seq is ideally suited for variant identification and tracking.

We envision that One-Seq can be clinically implemented in one of two ways to enable highly scalable viral diagnostics (Fig. 7A). First (the “v1” method), One-Seq can be directly used in a clinical lab with pre-collected specimens (e.g. swab or saliva in transport media) to achieve extraction-free, highly scalable diagnostics. Alternatively (the “v2” method), patient specimens can be directly collected into purpose-designed collection tubes containing One-Seq reagents and uniquely barcoded RT primers, and pooled immediately after incubation at the testing facility. The latter implementation would allow an even higher degree of scalability, as it completely avoids any liquid handling step for individual samples (Fig. 7B), and reduces the logistic complexity from one that scales with the number of samples to one that is largely independent of the number of samples (i.e. from O(N) to O(1)).

**Figure 7.**
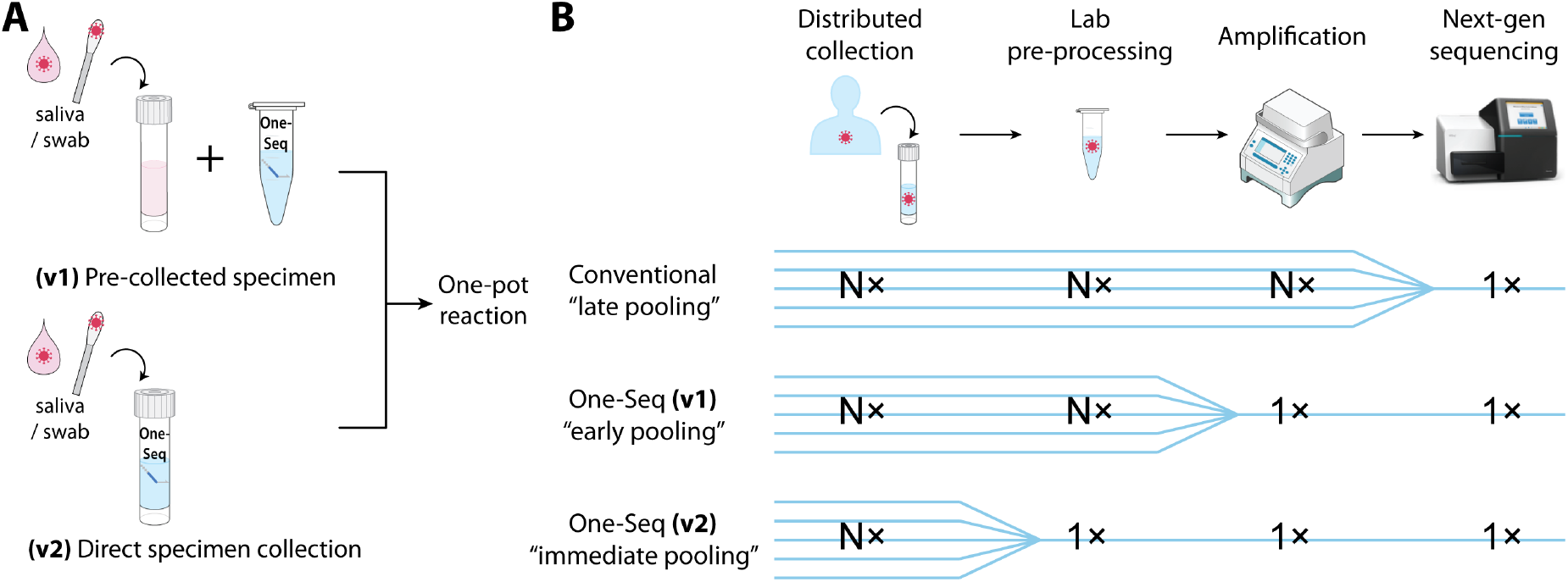
Potential clinical implementations for One-Seq. (**A**) Schematics for two clinical implementations: (**v1**) with pre-collected clinical specimen in viral transport medium, and (**v2**) with specimen collection directly into purpose-manufactured One-Seq collection tubes containing pre-assigned and uniquely identifiable sequence barcodes. (**B**) Compared with pre-collection (v1), direct collection (v2) completely avoids any liquid handling step and allows even higher scalability.

Finally, One-Seq is flexible in two important ways: it can be continually updated in a matter of days to include RT primers targeting emerging viral mutations as they appear, providing a real-time monitoring of viral evolution and transmission during an ongoing pandemic; and it can be targeted to detect any single-stranded RNA viruses of positive and negative sense, including the common cold, seasonal flu, hepatitis, dengue, Ebola, West Nile, Zika and more, potentially a number of them in a multiplexed manner. We envision that One-Seq would allow population-scale surveillance with a panel of viruses of special concern that could ultimately lead to the reporting of a “bio-weather map” for the early identification and tracking of emerging viral hotspots, and help prevent the next viral outbreak.

## Methods

### Clinical specimen and reference materials

All clinical specimen and saliva samples used in our study were de-identified. Remnant clinical nasopharyngeal swab samples were obtained from Boca Biolistics. The clinical specimens were not heat-inactivated prior to use, and all operations with clinical specimens were performed inside a biosafety cabinet (BSC) following BL2+ safety protocols. The use of clinical specimen was approved by the IRB at the Harvard Faculty of Medicine. SARS-CoV-2 inactivated virus standard materials were obtained from ATCC (VR-1986HK) or SeraCare (AccuPlex 0505-0168). In vitro transcribed viral N gene mRNA were prepared with Invitrogen MAXIscript T7 transcription kit (ThermoFisher, AM1312), following manufacturer’s protocol. The template DNA was prepared from N positive control plasmid (IDT, 10006625) with T7 promoter-containing primers, and purified from an agarose gel using QIAquick PCR purification kit (QIAGEN, 28104).

Acknowledgement for ATCC inactivated virus standard (VR-1986HK): The following reagent was deposited by the Centers for Disease Control and Prevention and obtained through BEI Resources, NIAID, NIH: Genomic RNA from SARS-Related Coronavirus 2, Isolate USA-WA1/2020, NR-52285

### Preparation of contrived specimens

For clinical limit of detection studies, we pooled confirmed COVID-19 negative remnant nasopharyngeal swab specimens purchased from Boca Biolistics (N=15). Pooled clinical samples were then spiked in with ATCC or SeraCare inactivated virus standard, or in vitro transcribed viral RNA, at various specified concentrations, pre-diluted into viral transport medium (VTM). VTM was prepared with 2% FBS (heat-inactivated at 56C for 30 min, Gibco 26140079), 1x Antibiotic-Antimycotic (Gibco, 15240096) and 11 mg/L phenol red, in 1x Hank’s balances salt solution (Gibco, 14025092). None of the contrived clinical samples were pre-heat-inactivated before one-pot reverse transcription step.

For reverse transcription efficiency studies, we pooled saliva specimen collected from COVID-19 negative donors, either with (N=4, “clean”) or without (N=9, “dirty) mouth rinsing before collection. Pooled saliva samples were then spiked with ATCC inactivated virus standard, or in vitro transcribed viral RNA, at specified concentrations, as above.

### Primer, barcode and sequencing construct designs

Reverse transcription primers were designed following these criteria: (i) Tm (calculated with IDT oligo analyzer, RNA-targeting primer) in range of 54-60C, strong 3’-end binding, and (ii) high sequence coverage of available SARS-CoV-2 genomes and low homology with SARS, MERS and related viral sequences. Furthermore, RT primers targeting mutation hotspots were design to be in close vicinity (within 5 nt) to the targeted loci, to avoid significantly increasing the sequencing runtime (Fig. 6A). Reverse primers for PCR are designed following these criteria: (i) Tm in range of 60-62C, weak 3’-end binding, and (ii) high sequence coverage of available SARS-CoV-2 genomes.

960 unique patient barcodes were designed by concatenating the i7 and i5 sequences and further expanding from IDT for Illumina Unique Dual Index set (4×96=384 pairs in total) (Fig. 3A).. The following sequences were inserted in between the sequence blocks: …AC…TG…AC… (4×96), …CA…CT…GA (4×96), …AC…AC…TG… (2×96). Such a design ensures a minimum Hamming distance of 12 between any two barcodes and avoids any homopolymer repeats longer than 3 nucleotides. 12 reverse PCR barcodes for batch pooling were selected from the set of IDT8 indices.

Sequencing constructs were designed using custom read primers and PCR adapters. Read primers were designed to be orthogonal to sequencing adapters and have Tm > 70C. A short PCR adapter sequence, which forms a part of the read 1 primer, was designed to allow for pooled amplification using a common forward primer and also compatible with the protector strand. A detailed illustration of the sequencing construct including example sequences are given in Fig. S2.

A full list of all primers, barcodes and adapters used in this study are provided in Tables S1-3 (Table S1: primers, adapters, batch barcodes, Table S2: 960 sample barcodes, Table S3: 96 selected sample barcodes).

### Synthetic positive control RNA

Positive control RNA was designed to start with the same RT primer with the N gene targeting primer N#1, and extended with 8 nt sequence distinct from the viral genome. Synthetic RNA was purchased from IDT, and spiked into all samples at a concentration of 10^4^-10^5^ copies/ul to provide positive control reads.

### One-pot sample processing reaction

One-pot sample reaction for viral lysis, reverse transcription and sample barcoding was performed with SuperScript IV reverse transcriptase (Thermo, 18090010) in manufacturer provided reaction buffer (without DTT), supplemented with 10% (v/v) murine RNAse inhibitor (New England Biolabs, M0314), 0.1% Triton X-100, 1x Antibiotic-Antimycotic (Gibco, 15240096), 0.5 mM EDTA, 5 mM DTT, cOmplete protease inhibitor cocktail (1 tablet into 13.3 ml, Sigma, 11873580001), 0.5 uM poly-A60 DNA oligonucleotide, 15 ug/ml E. coli tRNA (Sigma, 10109541001) and 10^4^-10^5^ copies/ul synthetic RNA for positive control, further added with 35-50% (v/v) equivalent of viral transport media or pooled clinical or saliva sample and 125 nM of barcoded RT primer (for each primer). For limit of detection studies, inactivated virus standard from ATCC or SeraCare was spiked into the one-pot reaction at specified concentrations. For barcode crosstalk studies, in vitro transcribed viral mRNA was used. For viral lysis and sample preservation studies, different subsets of above components were added to the reaction mix. For primer concentration studies, 25-500 nM of barcoded RT primers were used. For multiplexed sequencing samples, a master mix of above reaction mix without barcoded primer and contrived clinical sample was first prepared and aliquoted into a 96-well plate, then RT primers with unique barcodes and samples was added to each well.

One-pot reactions were assembled on ice-cold blocks. Once assembled, the reaction was incubated at 50C for 30 min, followed by inactivation at 95C for 5 min. For tests with contrived samples, incubation was performed in a closed-lid PCR thermocycler; for tests with clinical specimen, incubation was performed in a heat block, and followed by another inactivation session at 95C for 5 min in a closed-lid thermocycler once moved out of the BSC. For sample preservation studies, the assembled reaction was left at room temperature and covered for up to 24 hr before starting the 50C incubation.

### qPCR quantitation

For limit of detection studies for N#1 and N#2 primers, and RT quality control for clinical sample tests, qPCR was performed after the one-pot sample reaction. 0.5-1.0 ul one-pot reaction sample was added to 40 ul qPCR mix (40-80x dilution), containing Taq polymerase and standard buffer (New England Biolabs, M0273), 0.2 mM dNTP mix and CDC SARS-CoV-2 primer and probe set at 0.5 uM equivalent primer concentration (IDT RUO kit, 10006713). We observed formation of cloudy aggregation in certain clinical samples after the one-pot reaction. In such situation, to ensure adequate sample intake, the one-pot reactions were mixed with pipetting a few times before adding to the qPCR reaction. For limit of detection studies for variant targeting primers, qPCR was performed with dye-based readout, using Luna universal qPCR master mix (New England Biolabs, M3003) and 0.5 uM of both forward and reverse PCR primers.

qPCR samples were run on a Bio-Rad C1000 thermal cycler and CFX real-time PCR system for 50 cycles, and optionally with melt curve measurement for dye-based readout. Ct values were determined by manufacturer’s auto-thresholding function when possible. For preliminary clinical sensitivity studies, limit of detection (LoD) was determined to be the lowest viral spike-in concentration at which all 3/3 tests yielded a valid Ct value. For dye-based qPCR, results were interpreted with melt curve analysis instead of Ct values.

### Sample pooling and cDNA purification

One-pot reaction samples (20-80 ul each) were pooled by multichannel pipettes from 96-well plate to a single tube and immediately proceeded to cDNA purification using spin column (QIAquick PCR purification kit, QIAGEN 28104) or bead-based method (MagMax viral/pathogen nucleic acid isolation kit, Thermal A42352). We adapted the manufacturer’s protocols for large input sample volume and high sensitivity recovery. For column purification, sample was added multiple times to the same spin column. For bead purification, we used large 50 ml conical tubes and used centrifugation (3,000 rcf for 3 min) instead of magnetic attraction for effective collection of the beads. To ensure maximum recovery, we used all DNA low-bind tubes and pipette tips for this step. Purified cDNA library was supplemented with carrier DNA and RNA (poly-A60 oligonucleotide and E. coli tRNA) to further avoid sample loss on tube walls. For purification method comparison studies, we also compared QIAquick nucleotide removal kit (QIAGEN, 28304) and AmPure XP beads (Beckman Coulter, A63880), both following manufacturer’s protocols.

### Library amplification and quantitation

Pooled and purified cDNA library was amplified in a dUTP-incorporating PCR reaction, using Luna universal qPCR master mix (New England Biolabs, M3003), supplemented with UDG enzyme at 25 units/ml (New England Biolabs, M0372). For single-primer detection, 0.25 uM of both forward and reverse primers were used. For multi-primer detection with 4 primers, 0.5 uM of forward and 0.125 uM of each reverse primers were used. For multiplexed sequencing tests on clinical samples, 2 uM protector oligonucleotide was added. For protector concentration studies 0.5-5 uM protector was used. For barcode crosstalk studies, a mixture of 86 or 95 off-target barcoded RT primers was further supplemented into the reaction. For experiments containing positive controls (synthetic RNA, and human RPP control), PC libraries were amplified in separate reactions followed by independent normalization, to prevent inconclusive results due to lack of PC reads. Library amplification samples were run for 40-50 cycles with a custom-optimized thermocycling program: the first two cycles use a low annealing temperature (52-58C), and the rest use a high annealing temperature (68C).

The amplified library samples were within 200-260 bp range. Since non-specific amplification products can adversely affect loading concentration and sequencing quality, library quality was assessed on agarose gel and the desired band was purified using QIAqiuck PCR purification kit (QIAGEN, 28104). The purified library sample was then normalized using either Qubit or Agilent TapeStation before proceeding to sequencing run.

### Sequencing protocol

Sample libraries were sequenced on an Illumina MiSeq machine, at a loading concentration of 10 pM (for V2 Micro kit, 300-cycle, MS-103-1002) or 20 pM (for V3 kit, 150-cycle, MS-102-3001), supplemented with 15-20% Phi-X control v3 (Illumina, FC-110-3001). To avoid template carryover contamination between consecutive sequencing runs, we performed two template line washes (containing sodium hypochlorite solution, Sigma, 239305) between each run, following Illumina protocol.

Since our sequencing construct as well as barcodes were custom designed, we spiked in custom read primers into the sequencing kit following Illumina protocols (2 ul of 100 uM R1 custom read primer into well 12, and 2 ul of R2 primer into well 14). Sequencing was performed for 100+100 bases (for V2 Micro kit, 300-cycle) or 100+68 bases (for V3 kit, 150-cycle) with no indexing reads for developing the test, and can be shortened to 40-60 cycles for clinical use.

### Sequencing analysis

The bioinformatic analysis of sequencing results was performed in a few steps: FASTQ generation and adapter trimming (Illumina BaseSpace), sequence alignment (bowtie2), demultiplexing and read counting (custom scripts in MATLAB and Excel). Here sequence alignment was performed against sequences from one or multiple RT primers, allowing for ≤2 edit distance between library and sequencing read. In the case of viral sequencing and mutation identification, the reads were aligned against both original and mutated viral sequences, and the best matched genotype was reported. After alignment, each sample was identified using a combination of a front sample barcode, and a reverse batch barcode. All sequencing read counts were added by 1 to allow easy plotting. Our current analysis pipeline is not optimized and takes 20-30 min per run, however with further effort this method can be easily developed into a faster and more user-friendly analysis workflow.

### Analysis of barcode crosstalk and dynamic range

For barcode crosstalk studies with 1-10 high-load barcoded samples, supplemented with 86-95 off-target RT primers, after sequence alignment, the matched sequence counts for both groups of barcodes (on-target and off-target) were separated tallied. Read counts from the high-load samples were then normalized to 10^6^, then read counts from the off-target barcodes and relative level of crosstalk were determined.

### In silico analysis of primer specificity and inclusivity

In silico analysis for RT primer specificity and inclusivity was performed following FDA guideline (Molecular Diagnostic Template for Laboratories, version July 28, 2020). Specifically, inclusivity analysis was performed against all available SARS-CoV-2 genome sequences downloaded from NCBI (98,765 sequences, retrieved on Mar 9^th^, 2021), after excluding incomplete genomes (sequences with consecutive N’s and sequence fragments less than 20,000 nt in length). Specificity analysis was performed on Blastn against the recommended list of common respiratory flora and other viral pathogens (full list available in Table S5), using parameters optimized for detection of short, somewhat similar sequences.

### Confirmatory clinical sensitivity assay with multiplexed sequencing

Confirmatory clinical sensitivity studies were performed in pooled negative remnant clinical specimen background with different concentration of inactivated virus spike-in (ATCC) in a roughly 2x dilution series, based on results from pilot studies. All tests were performed with 96x multiplexed sample processing workflow. Each testing condition was repeated 20-22 times using high-quality, unique barcodes (i.e. not repeated 20-22 times with the same barcode) selected from barcode QC experiment. Each primer was tested multiple times with different batch barcode on the reverse side. Sequencing read threshold values were calculated using 3-σ formula (cut-off = mean + 3x stdev.) and reads obtained from negative control samples. The final limit of detection (LoD) for each target primer pair was determined using 95% detection rate cut-off (i.e. 19/20 or 21/22 detection) or 90% cut-off (when specified).

## Data Availability

Data reported in this study will be available upon request from the corresponding authors.
The nucleotide sequences of the SARS-CoV-2 genomes used in the primer inclusivity analysis are available from the NCBI database.

https://www.ncbi.nlm.nih.gov/sars-cov-2/

## References

1. B. J. Tromberg et al., Rapid Scaling Up of Covid-19 Diagnostic Testing in the United States - The NIH RADx Initiative. N Engl J Med 383, 1071–1077 (2020).

2. Y. C. Manabe, J. S. Sharfstein, K. Armstrong, The Need for More and Better Testing for COVID-19. JAMA, (2020).

3. M. Gandhi, D. S. Yokoe, D. V. Havlir, Asymptomatic Transmission, the Achilles’ Heel of Current Strategies to Control Covid-19. N Engl J Med 382, 2158–2160 (2020).

4. O. Vandenberg, D. Martiny, O. Rochas, A. van Belkum, Z. Kozlakidis, Considerations for diagnostic COVID-19 tests. Nat Rev Microbiol 19, 171–183 (2021).

5. J. R. Mascola, B. S. Graham, A. S. Fauci, SARS-CoV-2 Viral Variants-Tackling a Moving Target. JAMA, (2021).

6. M. J. MacKay et al., The COVID-19 XPRIZE and the need for scalable, fast, and widespread testing. Nat Biotechnol 38, 1021–1024 (2020).

7. M. N. Esbin et al., Overcoming the bottleneck to widespread testing: a rapid review of nucleic acid testing approaches for COVID-19 detection. RNA 26, 771–783 (2020).

8. R. Arnaout et al., SARS-CoV2 Testing: The Limit of Detection Matter. bioRxiv, (2020).

9. SARS-CoV-2 Reference Panel Comparative Data. U.S. Food and Drug Administration, (2020).

10. Roche Molecular Systems Inc., cobas SARS-CoV-2 Instructions for Use (Doc Rev. 5.0). U.S. Food and Drug Administration, (2020).

11. Broad Institute, COVID-19 Diagnostic Processing Dashboard. https://covid19-testing.broadinstitute.org/, (2021).

12. J. S. Bloom et al., Swab-Seq: A high-throughput platform for massively scaled up SARS-CoV-2 testing. medRxiv, (2020).

13. Illumina Inc., Illumina COVIDSeq Test Instructions for Use. U.S. Food and Drug Administration, (2020).

14. A. Ptasinska et al., Diagnostic accuracy of Loop mediated isothermal amplification coupled to Nanopore sequencing (LamPORE) for the detection of SARS-CoV-2 infection at scale in symptomatic and asymptomatic populations. medRxiv, (2020).

15. J. L. Schmid-Burgk et al., LAMP-Seq: Population-Scale COVID-19 Diagnostics Using Combinatorial Barcoding. bioRxiv, (2020).

16. Q. Wu et al., INSIGHT: A population-scale COVID-19 testing strategy combining point-of-care diagnosis with centralized high-throughput sequencing. Sci Adv 7, (2021).

17. R. Yelagandula et al., SARSeq, a robust and highly multiplexed NGS assay for parallel detection of SARS-CoV2 and other respiratory infections. medRxiv, (2020).

18. A. Hossain, A. C. Reis, S. Rahman, H. M. Salis, A Massively Parallel COVID-19 Diagnostic Assay for Simultaneous Testing of 19200 Patient Samples. Google Docs, (2020).

19. E. A. Bruce et al., Direct RT-qPCR detection of SARS-CoV-2 RNA from patient nasopharyngeal swabs without an RNA extraction step. PLoS Biol 18, e3000896 (2020).

20. Y. M. Bar-On, A. Flamholz, R. Phillips, R. Milo, SARS-CoV-2 (COVID-19) by the numbers. Elife 9, (2020).

21. Real-Time RT-PCR Panel for Detection, 2019-Novel Coronavirus - Instructions for Use. Center for Disease Control and Prevention, (2020).

22. I. Smyrlaki et al., Massive and rapid COVID-19 testing is feasible by extraction-free SARS-CoV-2 RT-PCR. Nat Commun 11, 4812 (2020).

23. S. Srivatsan et al., Preliminary support for a “dry swab, extraction free” protocol for SARS-CoV-2 testing via RT-qPCR. bioRxiv, (2020).

24. M. Kircher, S. Sawyer, M. Meyer, Double indexing overcomes inaccuracies in multiplex sequencing on the Illumina platform. Nucleic Acids Res 40, e3 (2012).

25. D. Y. Zhang, E. Winfree, Control of DNA strand displacement kinetics using toehold exchange. Journal of the American Chemical Society 131, 17303–17314 (2009).

26. PerkinElmer Inc., New Coronavirus Nucleic Acid Detection Kit Instructions for Use. U.S. Food and Drug Administration, (2020).

27. PerkinElmer Inc., PerkinElmer New Coronavirus Nucleic Acid Detection Kit Instructions for Use. U.S. Food and Drug Administration, (2020).

28. Hologic, Panther Fusion SARS-CoV-2 Assay Instructions for Use. U.S. Food and Drug Administration, (2020).

29. P. Horby et al., SAGE meeting report. (2021).

30. Y. Weisblum et al., Escape from neutralizing antibodies by SARS-CoV-2 spike protein variants. Elife 9, (2020).

31. W. F. Garcia-Beltran et al., Circulating SARS-CoV-2 variants escape neutralization by vaccine-induced humoral immunity. medRxiv, (2021).

