## Supplementary Info for "One-Seq: A Highly Scalable Sequencing-Based Diagnostic for SARS-CoV-2 and Other Single-Stranded Viruses"

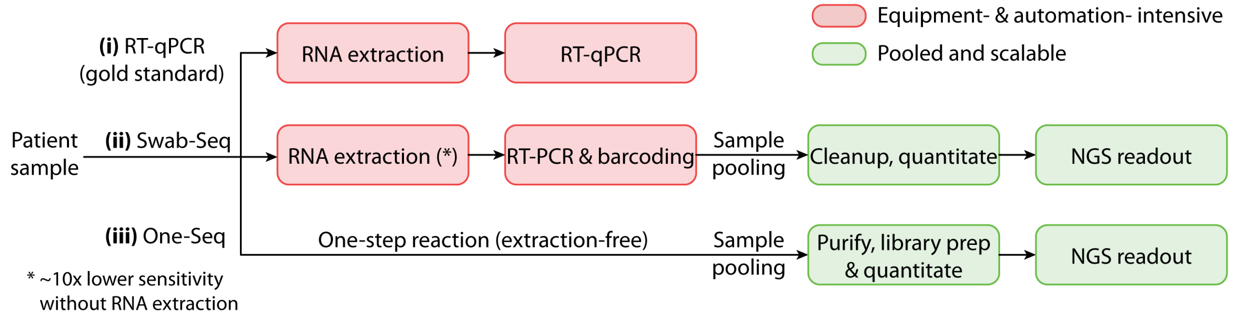

**Figure S1 Comparison of One-Seq workflow with other related methods**
Schematic comparison of sample processing workflow for (i) RT-qPCR (gold standard), (ii) Swab-Seq and (iii) One-Seq. One-Seq uses a one-step reaction to circumvent the need for RNA extraction and PCR amplification steps. Red blocks, sample processing steps that require high equipment usage and automation; green blocks, processing steps that are highly scalable.

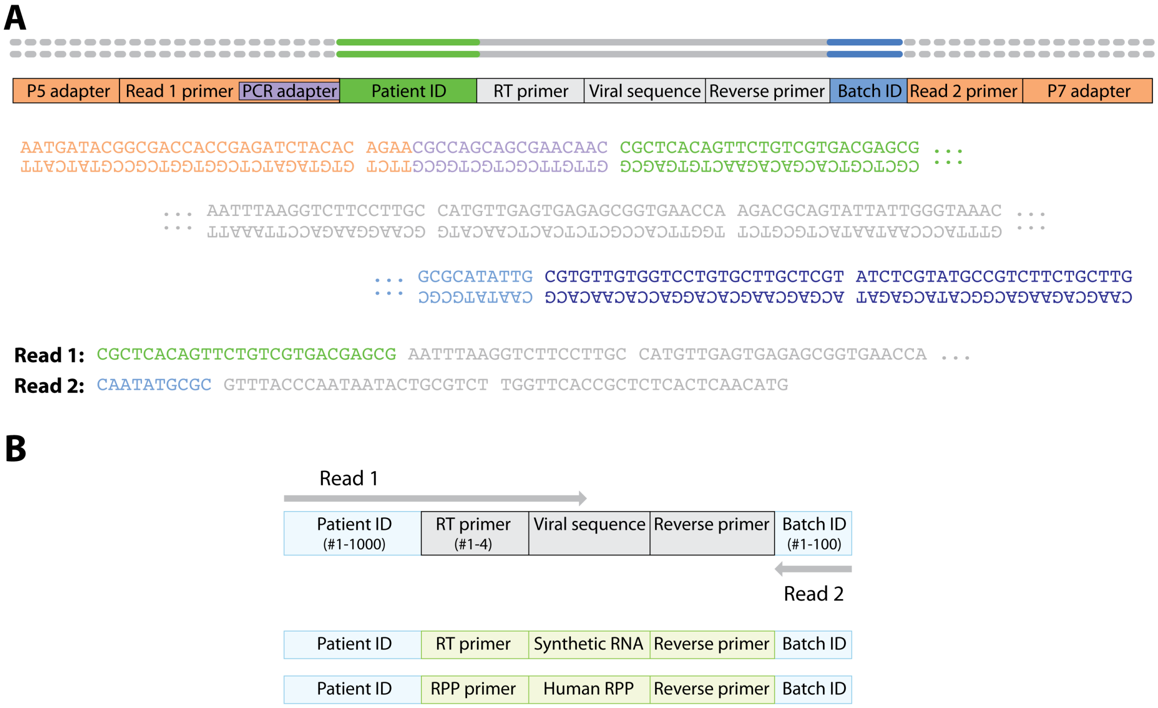

**Figure S2 One-Seq sequencing construct and read structure**
(**A**) Illustration of One-Seq sequnecing construct and example sequences. Each viral amplicon consists of a patient ID, RT primer, viral sequence, reverse primer, and batch ID. Sequences are illustrated with N#1 RT and PCR primers, barcode UDPS001 and S01. (**B**) Illustration of One-Seq sequencing read structure. Read 1 is used to decode patient ID (1000x), RT primer identity (4x) and amplicons from positive controls, read 2 is used to decode batch ID (100x).

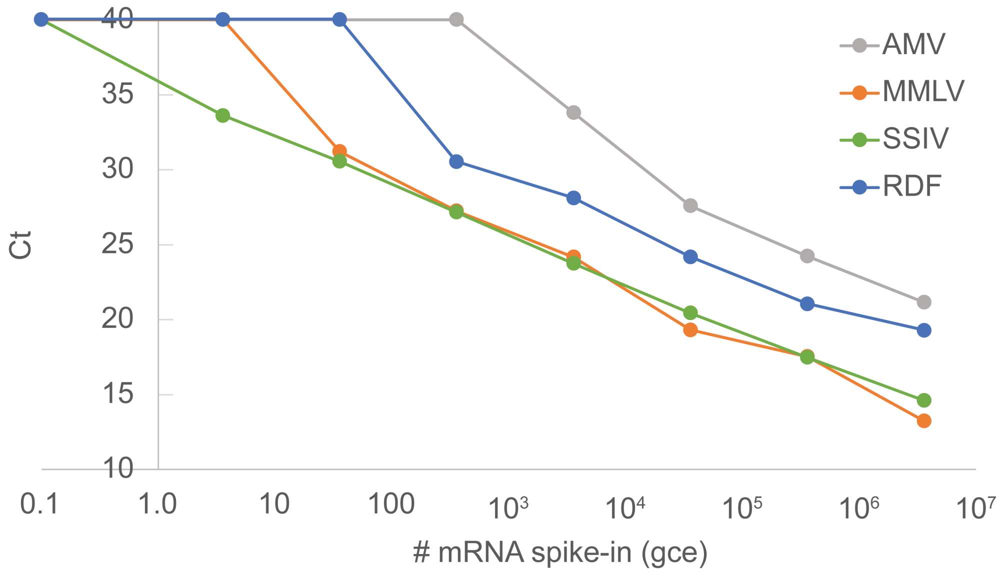

**Figure S3 Comparison of reverse transcriptase efficiency**
Reverse transcription (RT) efficiency of different RT enzymes were compared using two-step RT-qPCR and CDC's N gene primer and probe set (N1), in the presence of human saliva background (50% v/v) and RNAse inhibitor (Murine, 10% v/v). SSIV showed best RT efficiency in saliva containing samples, and detected 3 copies of mRNA spike-in. AMV, Avian Myeloblastosis Virus RT (New England Biolabs, M0277), MMLV, Moloney Murine Leukemia Virus RT (New England Biolabs, M0253), SSIV, SuperScript IV RT (ThermoFisher, 18090010), RDF, RapiDxFire (Lucigen, 30250).

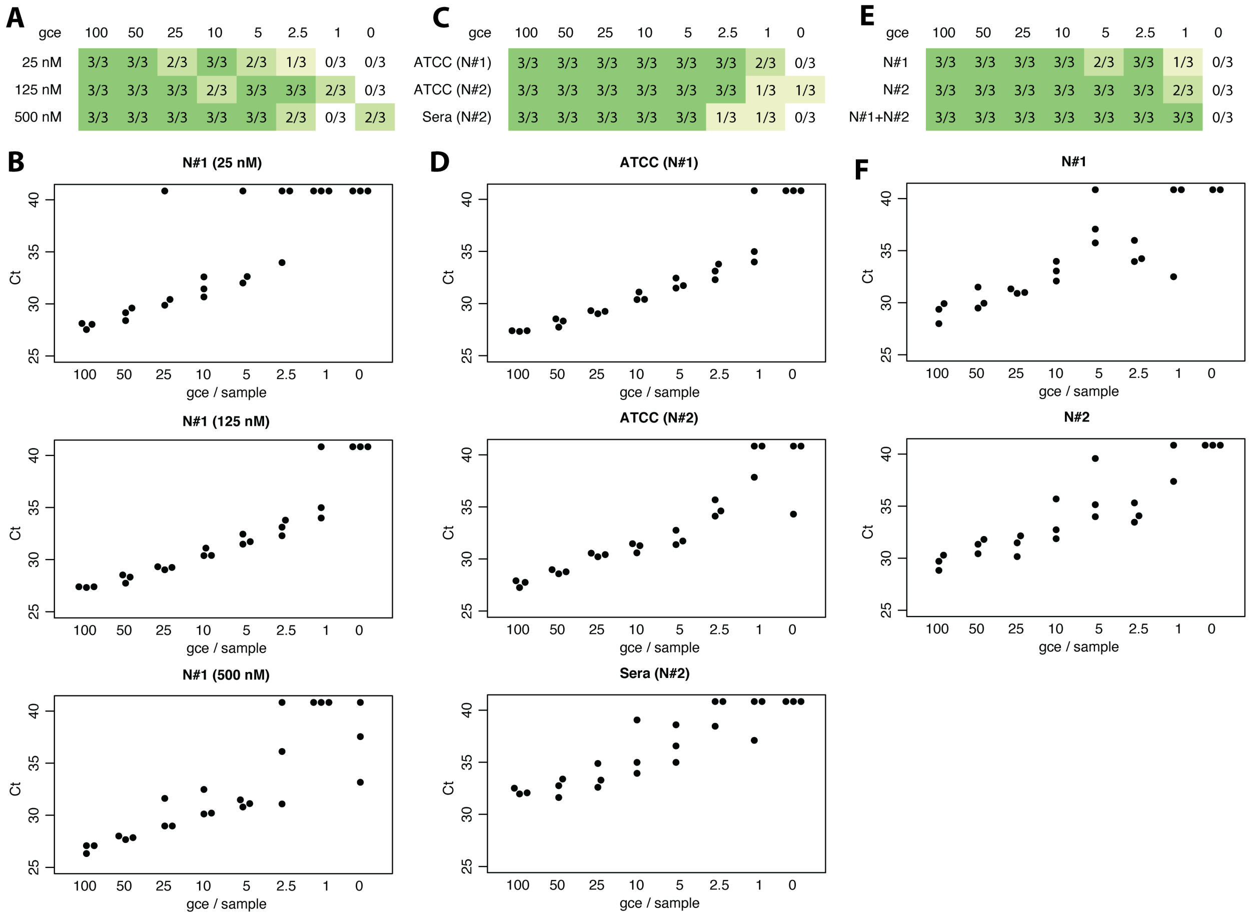

**Figure S4 Ct and limit of detection data for tests in Fig. 2C-E**
(**A-B**) Ct and limit of detection (LoD) data for main Fig. 2C, effect of RT primer concentration. (**C-D**) Ct and LoD data for main Fig. 2D, validation using different virus standard materials. (**E-F**) Ct and LoD data for main Fig. 2E, effect of multi-primer detection. (**A,C,E**) Limit of detection (LoD) determination. (**B,D,F**) Raw Ct data plots, each condition was repeated three times.

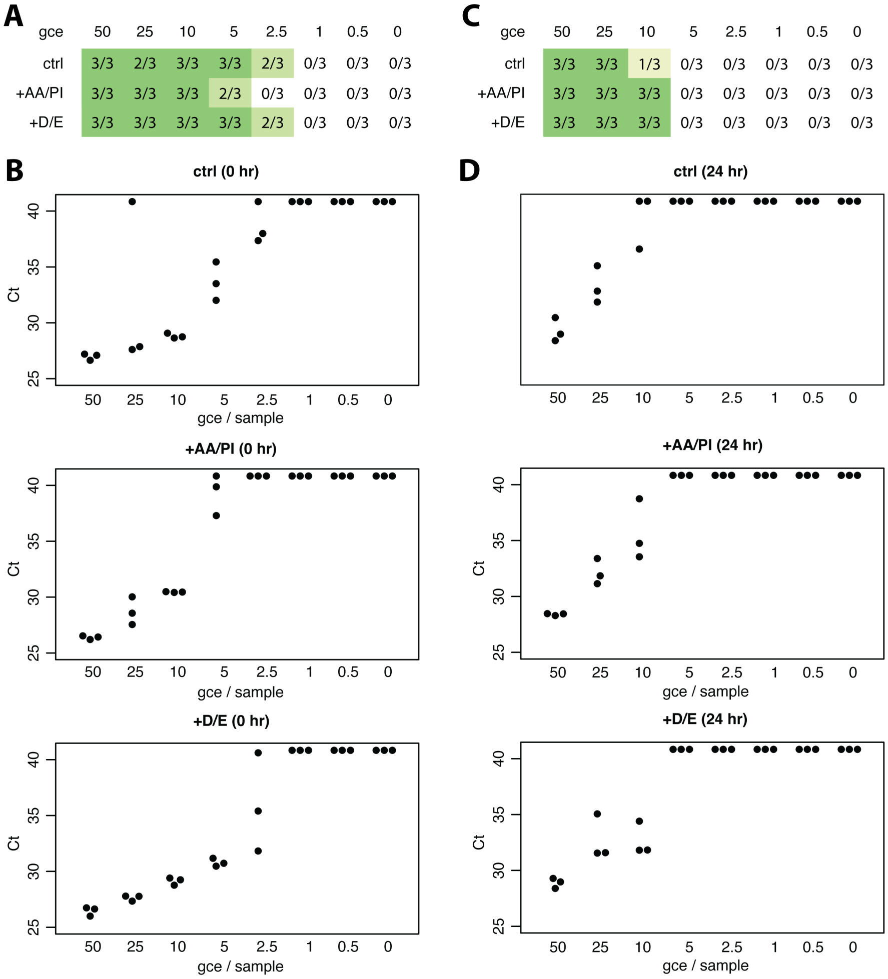

**Figure S5 Ct and limit of detection data for tests in Fig. 2F, effect of different sample preservatory buffers**

(**A,C**) Limit of detection (LoD) determination. (**B,D**) Raw Ct data plots, each condition was repeated three times.

**A**

| gce | 100 | 50 | 25 | 10 | 5 | 2.5 | 1 | 0 |
|---|---|---|---|---|---|---|---|---|
| ctrl | 3/3 | 2/3 | 3/3 | 3/3 | 3/3 | 3/3 | 2/3 | 1/3 |
| +AA/PI | 3/3 | 3/3 | 3/3 | 2/3 | 3/3 | 3/3 | 3/3 | 0/3 |
| +D/E | 3/3 | 3/3 | 3/3 | 3/3 | 3/3 | 3/3 | 2/3 | 0/3 |

**C**

| gce | 100 | 50 | 25 | 10 | 5 | 2.5 | 1 | 0 |
|---|---|---|---|---|---|---|---|---|
| ctrl | 3/3 | 3/3 | 3/3 | 3/3 | 3/3 | 2/3 | 2/3 | 0/3 |
| +AA/PI | 3/3 | 3/3 | 3/3 | 3/3 | 3/3 | 3/3 | 1/3 | 0/3 |
| +D/E | 1/3 | 0/3 | 0/3 | 0/3 | 0/3 | 0/3 | 0/3 | 0/3 |

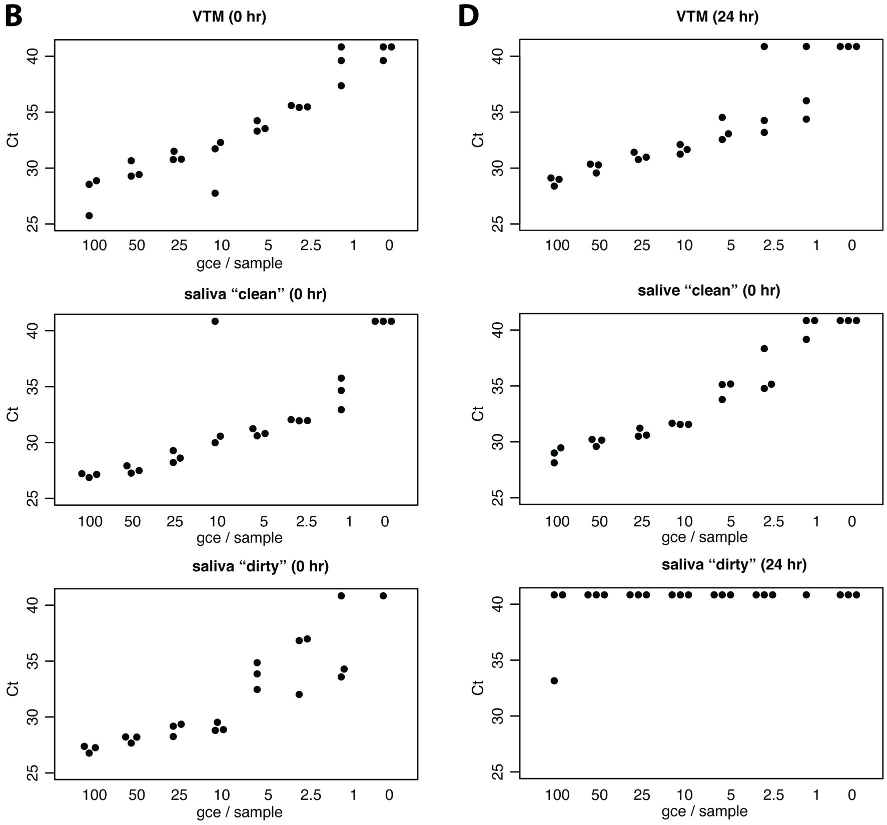

**Figure S6 Ct and limit of detection data for tests in Fig. 2G, effect of sample preservatory buffers in VTM and saliva samples**

(**A,C**) Limit of detection (LoD) determination. (**B,D**) Raw Ct data plots, each condition was repeated three times.

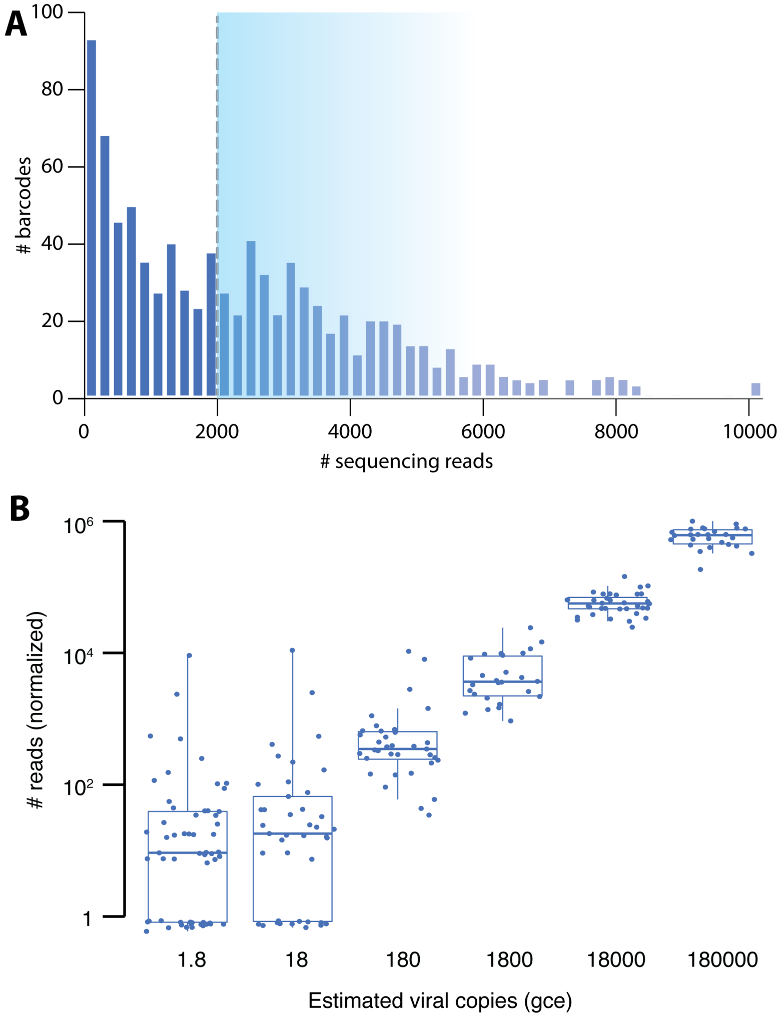

**Figure S7 960x barcode QC and selection**

(**A**) Distribution of sequencing reads from all 960x sample barcodes, barcodes with reads above median were selected for later tests. (**B**) Preliminary linearity and dynamic range test with 200x selected barcodes. Sequencing reads showed linear response at higher viral load conditions and dynamic range of ~$10^4$.

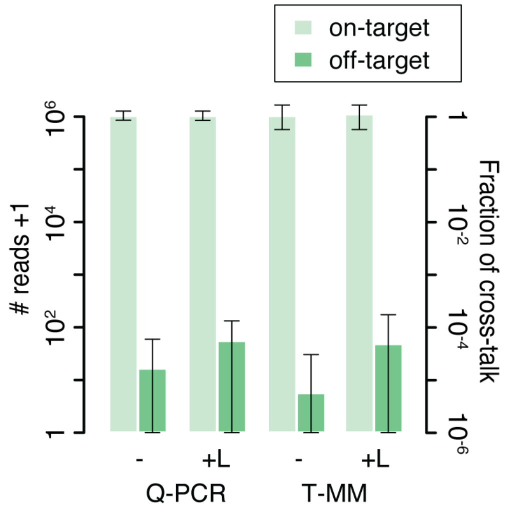

**Figure S8 Barcode crosstalk and dynamic range test in 10-plex settings**

Barcode crosstalk and dynamic range was tested with 10 high-load samples, in the presence of ~86x off-target primers, amplified in the presence of protector strand, and assayed by sequencing. Four conditions were tested, using two different cDNA purification methods (Q-PCR and T-MM) and with or without supplementation of extra off-target primers (-, without supplementation, +L, with low amount supplementation). Reads were normalised by on-target samples (average) to $10^6$ reads per barcode. Q-PCR, QIAquick PCR purification kit (QIAGEN), T-MM, MagMax viral/pathogen nucleic acid isolation kit (ThermoFisher).

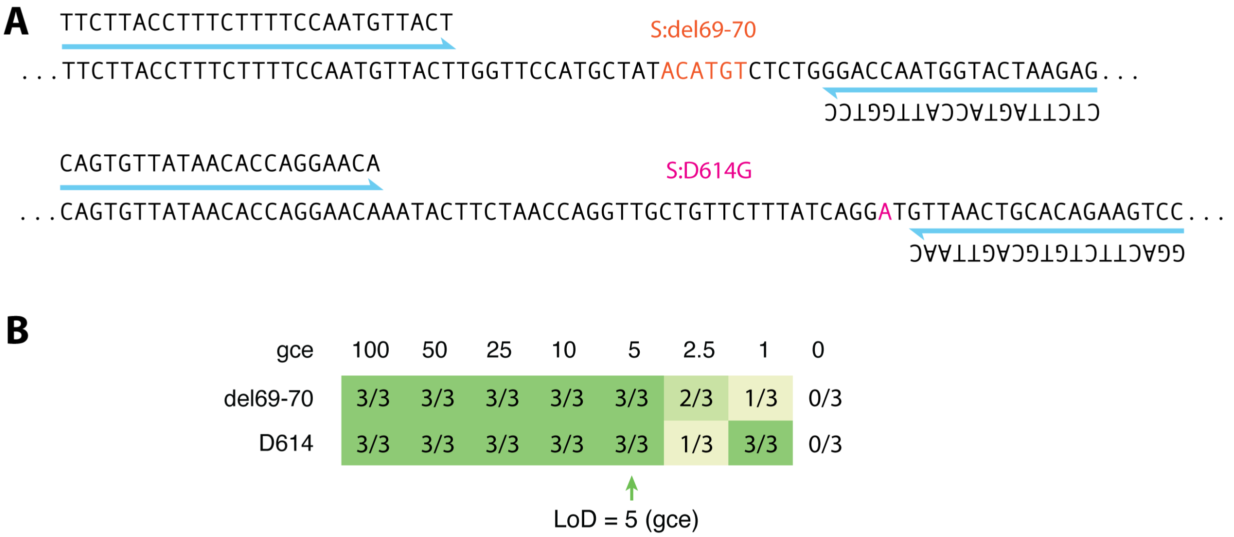

**Figure S9 Design of RT and PCR primers targeting viral hotspot mutations and RT sensitivity test**

(**A**) Sequence design of RT and PCR primers targeting two SARS-CoV-2 hotspot mutations, S:del69-70 and S:D614G. Nucleotides affected by these mutations are coloured. (**B**) RT sensitivity assay by dye-based qPCR assay, using above primer sets. LoD determined to be 5 gce for both targets.

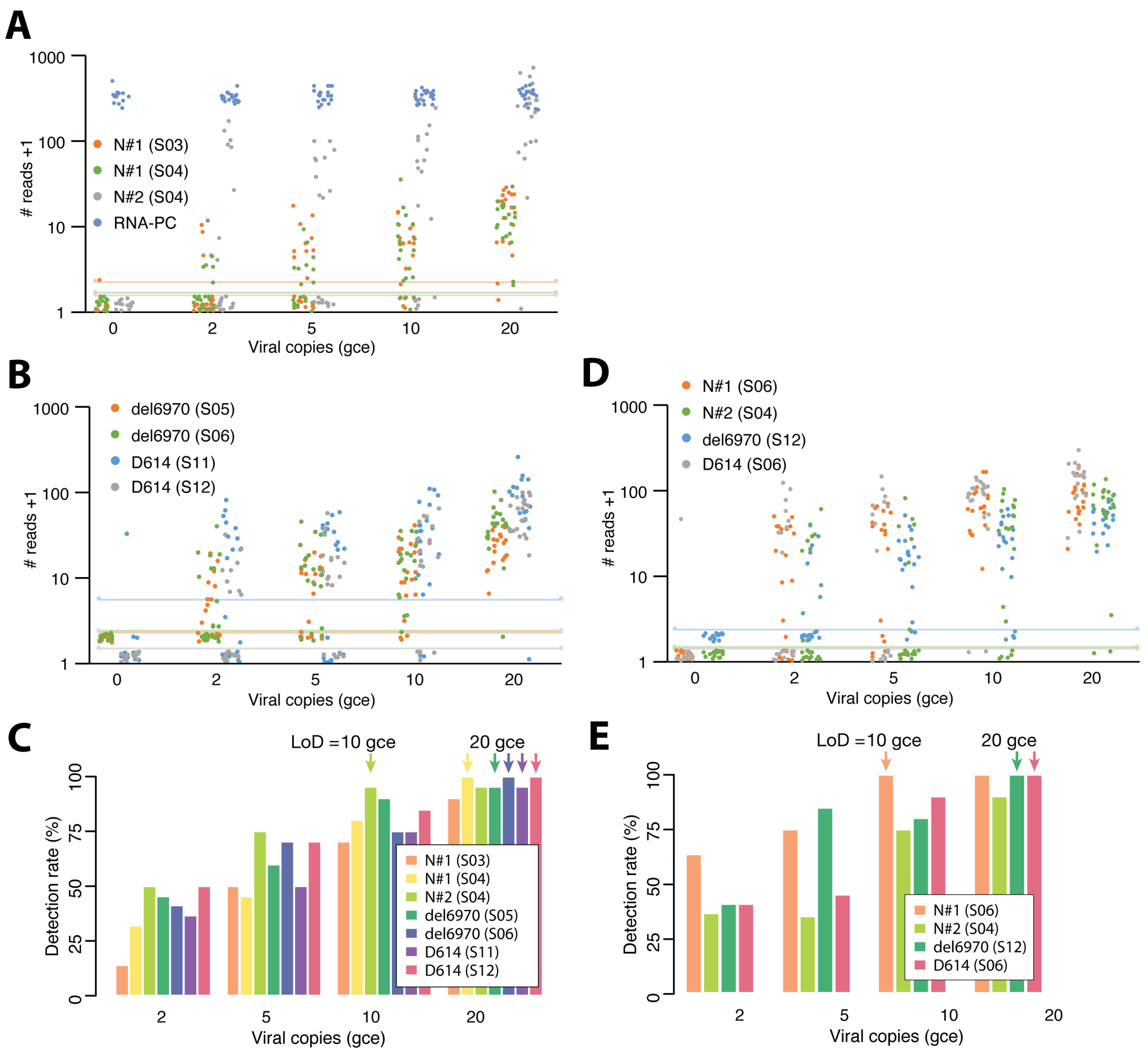

**Figure S10 Confirmatory clinical sensitivity studies in 96x multiplexed test**

Confirmaty clinical sensiticity studies were performed in pooled negative remnant clinical specimen background with different concentration of inactivated virus spike-in. All tests were performed with 96x multiplexed sample processing workflow. Each testing condition was repeated 20-22 times using unique barcodes (i.e. **not** repeated 20-22 times with the same barcode). Each primer was tested multiple times with different batch barcode on the reverse side. LoD was determined using 95% detection rate criteria (i.e. 19/20 detection). (**A-C**) Confirmatory clinical sensitivity studies for single-primer detection. (**D-E**) Confirmatory clinical sensitivity studies for multi-primer detection. (**A,B,D**) Each test condition was repeated 20-22 times with unique barcodes. Plots showing sequencing reads for each barcode and each test condition. Solid lines indicate 3-σ threshold values. S number indicates batch barcode. (**C,E**) Detection rate at different viral load conditions and LoD values determined for each primer.

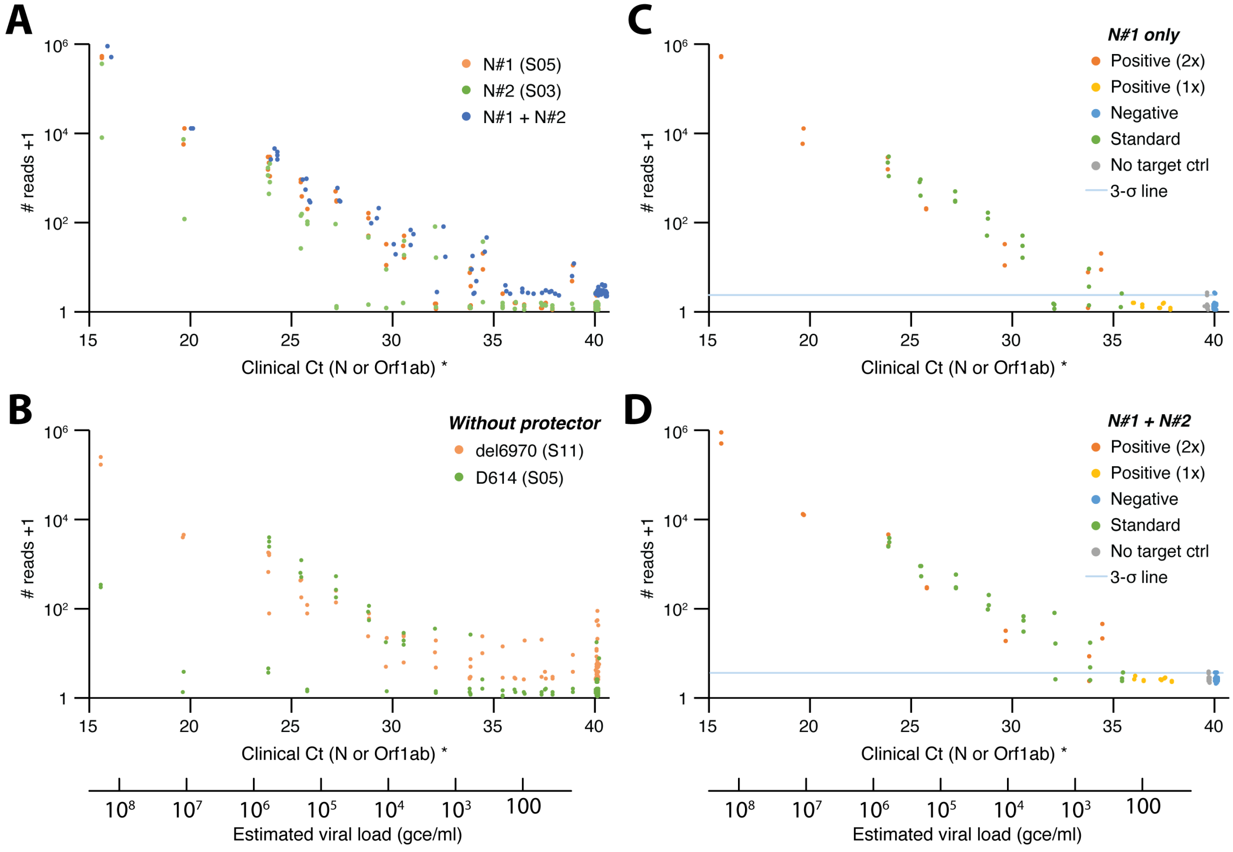

**Figure S11 Raw sequencing reads and breakdown for multi-primer clinical sample test in 96x multiplexed test**

Raw sequencing reads were plotted against clinical Ct values for N gene or Orf1ab gene (if N gene was not detected). (**A,B**) Sequencing read scatter plot for all samples, including clinical samples, standards and negative controls, and all four viral targeting primers. S number indicates batch barcode. Note that del6970 and D614 targets were amplified in the absence of protector strand, and shows a limited dynamic range as a result. (**C,D**) Breakdown of sequencing read for N#1 and N#2 primers, individually (**C**) and summed together (**D**). Positive (2x) refers to samples for which clinical RT-qPCR test returned positive results for both N and orf1ab amplicons, and positive (1x) refers to samples for which only one of the two amplicons were clinically detected (and Ct>36).

**Table S1** One-Seq primers, adapters, batch barcodes and protector strands

All Tm values were calculated using IDT oligo analyzer (https://www.idtdna.com/calc/analyzer), with qPCR default parameters.

| Name | Type | Sequence | Tm |
|---|---|---|---|
| N#1_RT | RT primer | AATTTAAGGTCTTCCTTGC | 53.8C (RNA) |
| N#1_PCR | Reverse PCR primer | GTTTACCCAATAATACTGCGTCT | 60.8C |
| N#2_RT | RT primer | TGTGTAGGTCAACCACG | 53.7C (RNA) |
| N#2_PCR | Reverse PCR primer | CAGACAAGGAACTGATTACAAACA | 61.5C |
| del6970_RT | RT primer | CTCTTAGTACCATTGGTCC | 61.4C (RNA) |
| del6970_PCR | Reverse PCR primer | TTCTTACCTTTCTTTTCCAATGTTACT | 62.0C |
| D614_RT | RT primer | GGACTTCTGTGCAGTTAAC | 56.5C (RNA) |
| D614_PCR | Reverse PCR primer | CAGTGTTATAACACCAGGAACA | 60.3C |
| RNA_PC | Synthetic RNA control | CCAAGGTTTACCCAATAATACTGCTGAGGTTGTCAC CGCTCTCACGACCACGTGCAAGGAAGACCTTAAATT | |
| RNA_PC_PCR | Reverse PCR primer | CCAATAATACTGCTGAGGTTGT | 60.5C |
| P5xs | Short PCR adapter | CGCCAGCAGCGAACAA | 62.8C |
| P5xe | P5 side adapter and common PCR primer | AATGATACGGCGACCACCGA GATCTACAC AGAACGCCAGCAGCGAACAA | 78.4C |
| P5xr | Read 1 primer | CGA GATCTACAC AGAACGCCAGCAGCG | 70.9C |
| P7y | P7 side adapter | CAAGCAGAAGACGGCATACGAGAT ACGAGCAAGCACAGG ACCACAACACG | 77.4C |
| P7yr | Read 2 primer | ACGAGCAAGCACAGG ACCACAACACG | 71.7C |
| S01 | Batch barcode | TGGTACAG | |
| S02 | Batch barcode | AACCGTTC | |
| S03 | Batch barcode | TAACCGGT | |
| S04 | Batch barcode | GAACATCG | |
| S05 | Batch barcode | CCTTGTAG | |
| S06 | Batch barcode | TCAGGCTT | |
| S07 | Batch barcode | GTTCTCGT | |
| S08 | Batch barcode | AGAACGAG | |
| S09 | Batch barcode | TGCTTCCA | |
| S10 | Batch barcode | CTTCGACT | |
| S11 | Batch barcode | CACCTGTT | |
| S12 | Batch barcode | TGGTACAG | |
| N#1_prot | Protector strand | AATTTAAGGTCTTCCTTGC CATGTTGAGTGAGAG CGGTGAACCAAGACG /3InvdT/ | |
| N#2_prot | Protector strand | TGTGTAGGTCAACCACG TTCCCGAAGGTGTGA CTTCCATGCCAATGC /3InvdT/ | |

**Table S2** 960x unique sample barcodes (Barcode IDs: UDPX001-960)

(See separate Excel table for barcode sequences.)

**Table S3** 96x selected sample barcodes (Barcode IDs: UDPS001-096)

| UDPS ID | UDPX ID | Barcode Sequence |
|---|---|---|
| UDPS001 | UDPX100 | TCTCCACATTGATGGCACTACACAAC |
| UDPS002 | UDPX101 | ACATGACCATATTGTGGTGACCCTGG |
| UDPS003 | UDPX102 | CAGGCACGCCATTGTCCACACGGCCT |
| UDPS004 | UDPX106 | TTCTAACCAGAATGGAGCGACCAATA |
| UDPS005 | UDPX107 | TATTGACCGTTCTGATCTTACACTGT |
| UDPS006 | UDPX330 | GAAGCACTAGCTTGTAATAACCGGAG |
| UDPS007 | UDPX196 | CCATCACCACGCTGTTGACACCAATG |
| UDPS008 | UDPX197 | ACAACACCAGGATGCTGACACCGGCA |
| UDPS009 | UDPX354 | ACACAACGCGCTTGCGTTCACGGAAC |
| UDPS010 | UDPX202 | AGTTAACTCACATGCAGGCACGCCAT |
| UDPS011 | UDPX203 | GTAGCACATACTTGTTAATACAGACC |
| UDPS012 | UDPX204 | CTTCAACGTTACTGGGAGTACCGCGA |
| UDPS013 | UDPX112 | GTAGCACCATCATGCCAACACAACAT |
| UDPS014 | UDPX113 | CTTGTACAATTCTGACCGGACCTCAG |
| UDPS015 | UDPX114 | TCCAAACTTCTATGGTTAAACTCTGA |
| UDPS016 | UDPX118 | GAACAACAGTATTGTAACCACGCCGA |
| UDPS017 | UDPX119 | AATTGACGCGGATGCTCCGACTGCTG |
| UDPS018 | UDPX120 | GGCCTACGTCCTTGCATTCACCAGCT |
| UDPS019 | UDPX208 | TTATCACCGATCTGGTATCACGGCCG |
| UDPS020 | UDPX209 | ATAGTACCTAGCTGAATACACGACAT |
| UDPS021 | UDPX378 | AGAACACCGCGGTGGGAGTACAGATT |
| UDPS022 | UDPX214 | TCGCGACTATAATGGCCGTACCTGTT |
| UDPS023 | UDPX215 | ATTCTACAAGCGTGCAGAGACTGATA |
| UDPS024 | UDPX216 | AGCGCACTTCGGTGTGCTAACACTAT |
| UDPS025 | UDPX292 | TGGCAACATATTTGCGGTGACACACC |
| UDPS026 | UDPX125 | TCCACACGGCCTTGCTCGTACGCGTT |
| UDPS027 | UDPX294 | CGTGTACATCTTTGTGTGCACTAACA |
| UDPS028 | UDPX130 | TCTTGACGCTATTGAGTGCACCACTG |
| UDPS029 | UDPX131 | TCACAACCCGAATGGAACAACAGTAT |
| UDPS030 | UDPX132 | AACGTACTACATTGACGATACTGCTG |
| UDPS031 | UDPX220 | GCGTAACCTTAGTGAACATACACCTA |
| UDPS032 | UDPX221 | TACCGACAACTATGCCATGACTGTAG |
| UDPS033 | UDPX312 | ACTTCACCTAGCTGTCTCGACGACGA |
| UDPS034 | UDPX376 | GGACGACTCTTGTGAGTATACACGGA |
| UDPS035 | UDPX227 | AATATACGAAGCTGCGTTCACAGCCT |
| UDPS036 | UDPX228 | TAGCGACCTAGTTGTTACTACTCCTC |
| UDPS037 | UDPX136 | TGGTGACCCTGGTGACGCTACAATTA |
| UDPS038 | UDPX137 | TAGGAACACCGGTGTATATACTCGAG |
| UDPS039 | UDPX138 | AATATACTGGCCTGCGGTCACCGATA |
| UDPS040 | UDPX142 | CAGTAACGTTGTTGAGTTAACTCACA |
| UDPS041 | UDPX143 | TTCATACCCAACTGTTCCAACGGTAA |
| UDPS042 | UDPX144 | CAATTACGGATTTGCATGTACAGAGG |
| UDPS043 | UDPX328 | ACTAGACCCGTGTGGGTGTACACAAG |
| UDPS044 | UDPX353 | GTGTGACATATCTGGACTAACTATGT |
| UDPS045 | UDPX234 | GTCACACCACAGTGGACAGACACAGG |
| UDPS046 | UDPX238 | TGGTTACAAGAATGTCTACACATACC |
| UDPS047 | UDPX239 | ACTCTACTCCTTTGCACGTACTAGGC |
| UDPS048 | UDPX240 | GTCTCACCTTCCTGTGGTGACAGTCT |
| UDPS049 | UDPX148 | CTATAACCGCGGTGTAGGAACACCGG |

| | | |
|---|---|---|
| UDPS050 | UDPX149 | ATTCAACGAATCTGAGCGGACTGGAC |
| UDPS051 | UDPX150 | GTATTACCTCTATGTATAGACATTCG |
| UDPS052 | UDPX154 | AACTCACCGAACTGCGCCTACTCTGA |
| UDPS053 | UDPX341 | ATCGTACCGCTCTGCGTATACAATCA |
| UDPS054 | UDPX156 | TGAATACATTGCTGGGCGCACCAATT |
| UDPS055 | UDPX340 | GACTGACGTTGCTGCACCGACAGGAA |
| UDPS056 | UDPX245 | TGGTCACTAGTGTGACTGCACCTTAT |
| UDPS057 | UDPX324 | CACCTACCTTGGTGAACGAACGCCAG |
| UDPS058 | UDPX322 | TGCCTACACGAGTGTCATCACCTCTT |
| UDPS059 | UDPX251 | ATTAAACTACGCTGTGCGGACTGTTG |
| UDPS060 | UDPX252 | TAGTCACACAACTGACATAACACGGA |
| UDPS061 | UDPX160 | GCTCCACGTCACTGTGGCGACGTCCA |
| UDPS062 | UDPX305 | TGTGAACTGTATTGTCTCCACATTGA |
| UDPS063 | UDPX162 | ACACCACGTTAATGTCCTGACACCGT |
| UDPS064 | UDPX166 | ATCGCACATATGTGCCTTGACAACGG |
| UDPS065 | UDPX167 | ATCATACAGGCTTGCACCAACCCTAC |
| UDPS066 | UDPX342 | GGTGCACGTTCGTGATGACACAGAAC |
| UDPS067 | UDPX364 | AGCATACTAACTTGACTGCACAGCCG |
| UDPS068 | UDPX257 | AGATTACGTTACTGCTCTGACTATAC |
| UDPS069 | UDPX336 | TTGAGACCCTAATGTCGAAACTGGAA |
| UDPS070 | UDPX262 | CATGGACTCTAATGTCCTAACGGAAG |
| UDPS071 | UDPX377 | GTTGTACACTCATGACGCTACTGGAC |
| UDPS072 | UDPX264 | GCCGAACCAAGATGCCACCACTGTGT |
| UDPS073 | UDPX172 | CAACCACGGAGGTGCATAAACCACCA |
| UDPS074 | UDPX173 | AGCGGACTGGACTGTCCTAACTTAGC |
| UDPS075 | UDPX318 | TACTAACACACATGTACTCACTGTTA |
| UDPS076 | UDPX178 | CGCGAACCGATCTGCTGGAACTATGT |
| UDPS077 | UDPX179 | GCCTCACGGATATGGGCCAACATAAG |
| UDPS078 | UDPX180 | TGAGAACCAGCGTGATTACACTCACC |
| UDPS079 | UDPX268 | AGACTACCTCTTTGTACGAACATCTT |
| UDPS080 | UDPX269 | GCTCGACCCTACTGTAGGAACGCGCA |
| UDPS081 | UDPX348 | GTGGCACTGGTTTGGCCACACAGCAC |
| UDPS082 | UDPX274 | AATAGACGCCTCTGCTAGTACCCGGA |
| UDPS083 | UDPX275 | CCGCTACTAGCTTGATTAAACTACGC |
| UDPS084 | UDPX276 | TCCTAACGGAAGTGCCTAGACAGTAT |
| UDPS085 | UDPX304 | AACGGACAGCGGTGTACGGACCGAAG |
| UDPS086 | UDPX185 | GTTGGACACCGTTGTCTTAACCATCA |
| UDPS087 | UDPX186 | ACCAAACGTTACTGCGCCAACTACCT |
| UDPS088 | UDPX190 | GATTAACAGGTGTGTTAGGACATAGA |
| UDPS089 | UDPX191 | CAACAACTTCAATGCGCAAACTCTAG |
| UDPS090 | UDPX192 | GTGTTACACCGGTGGAGTTACGTACT |
| UDPS091 | UDPX280 | CACTTACAATCTTGCACCTACCTTGG |
| UDPS092 | UDPX365 | ATTACACTCACCTGAACATACCTAGT |
| UDPS093 | UDPX282 | GGCGAACATTCTTGCGGCAACAGCTC |
| UDPS094 | UDPX286 | CCTTAACCTATGTGACCAAACGTTAC |
| UDPS095 | UDPX323 | ACTAGACAACTTTGGGTAAACGATAA |
| UDPS096 | UDPX288 | TAATCACGGTACTGAACTAACACGTT |

**Table S4** Inclusivity analysis of primers used

| Sequence homology | N#1 | | N#2 | | del6970 | | D614 | |
|---|---|---|---|---|---|---|---|---|
| | RT | PCR | RT | PCR | RT | PCR | RT | PCR |
| Exact | 99.7% | 99.8% | 97.8% | 99.5% | 99.6% | 99.4% | 99.6% | 99.9% |
| ≤1 nt mismatch | 100.0% | 100.0% | 100.0% | 100.0% | 100.0% | 100.0% | 100.0% | 100.0% |

**Table S5** List of organisms and taxonomy ID used for cross-reactivity analysis

| Organism | Taxonomy ID |
|---|---|
| Human adenovirus C1 | 10533 |
| Human adenovirus A | 129875 |
| adenovirus Ad5 | 28285 |
| Human metapneumovirus | 162145 |
| Human parainfluenza virus 1 | 12730 |
| Human parainfluenza virus 2 | 1979160 |
| Human parainfluenza virus 3 | 11216 |
| Human parainfluenza virus 4 | 11203 |
| Human adenovirus 7 | 10519 |
| Influenza virus type A | 11320 |
| Influenza virus type B | 11520 |
| Human enterovirus EV68 | 42789 |
| Human respiratory syncytial virus | 11250 |
| Rhinovirus | 12059 |
| Chlamydia pneumoniae | 83558 |
| Haemophilus influenzae | 727 |
| Legionella pneumophila | 446 |
| Mycobacterium tuberculosis | 1773 |
| Streptococcus pneumoniae | 1313 |
| Streptococcus pyogenes | 1314 |
| Bordetella pertussis | 520 |
| Mycoplasma pneumoniae | 2104 |
| Pneumocystis jirovecii | 42068 |
| Candida albicans | 5476 |
| Pseudomonas aeruginosa | 287 |
| Staphylococcus epidermidis | 1282 |
| Streptococcus salivarius | 1304 |
| Human coronavirus (STRAIN 229E) | 11137 |
| Human coronavirus (strain OC43) | 31631 |
| Human coronavirus NL63 | 277944 |
| Human coronavirus HKU1 | 290028 |
| MERS | 1335626 |
| HCoV-SARS | 694009 |

**Table S6** Breakdown of One-Seq processing times

| One-Seq workflow | | Processing time | | |
|---|---|---|---|---|
| | | MiSeq | NextSeq 550 | NovaSeq 6000 |
| **(1) Diagnostic workflow** | | | | |
| Sample incubation (one-pot reaction and inactivation) | | 40 min | | |
| Sample pooling and cDNA purification | | 60 min | | |
| Library amplification | | 90 min | | |
| Purification and quantitation | | 60 min | | |
| Sequencing (diagnostics only) | Cluster generation | 60 min | 150 min | 130 min |
| | Patient barcode (R1, 26 nt) | 120 min | 120 min | 180 min |
| | RT primer ID (R1, 5 nt) | 20 min | 20 min | 30 min |
| | (subtotal) | 200 min | 290 min | 340 min |
| **(2) Optional – Batch pooling** | | | | |
| Sequencing (batch pooling) | Paired-end turn-around | 30 min | 60 min | 50 min |
| | Batch barcode (R2, 10 nt) | 45 min | 45 min | 70 min |
| | (subtotal) | 75 min | 105 min | 120 min |
| **(3) Optional – Variant identification** | | | | |
| Sequencing (variant ID) | RT primer and mutation hotspot (R1, 20 nt) | 100 min | 100 min | 150 min |

**Table S7** Breakdown of One-Seq reagent cost

| Component | Current cost* (off-the-shelf) | Estimated future cost** | Product and manufacturer |
|---|---|---|---|
| RNAse inhibitor | $ 2.5 | $ 0.63 | Murine (New England Biolabs, M0314) |
| RT enzyme | $ 2.6 | $ 0.66 | SuperScript IV (ThermoFisher, 18090010) |
| Chemicals, oligonucleotides, and other additives | $ <0.2 | $ <0.1 | (various) |
| *Total* | *$ 5.3* | *$ 1.39* | |

* All costs are estimated for 10 ul patient sample input. For 5 ul patient sample input, all costs are reduced by a half.

** Enzyme costs can be significantly reduced when mass produced, estimated as 25% of current off-the-shelf cost.
